# A comprehensive workflow for target adaptive sampling long-read sequencing applied to hereditary cancer patient genomes

**DOI:** 10.1101/2023.05.30.23289318

**Authors:** Wataru Nakamura, Makoto Hirata, Satoyo Oda, Kenichi Chiba, Ai Okada, Raúl Nicolás Mateos, Masahiro Sugawa, Naoko Iida, Mineko Ushiama, Noriko Tanabe, Hiromi Sakamoto, Yosuke Kawai, Katsushi Tokunaga, NCBN Controls WGS Consortium, Shinichi Tsujimoto, Norio Shiba, Shuichi Ito, Teruhiko Yoshida, Yuichi Shiraishi

## Abstract

Innovations in sequencing technology have led to the discovery of novel mutations that cause inherited diseases. However, many patients with suspected genetic diseases remain undiagnosed. Long-read sequencing technologies are expected to significantly improve the diagnostic rate by overcoming the limitations of short-read sequencing. In addition, Oxford Nanopore Technologies (ONT) offers a computationally-driven target enrichment technology, adaptive sampling, which enables intensive analysis of targeted gene regions at low cost. In this study, we developed an efficient computational workflow for target adaptive sampling long-read sequencing (TAS-LRS) and evaluated it through application to 33 genomes collected from suspected hereditary cancer patients. Our workflow can identify single nucleotide variants with nearly the same accuracy as the short-read platform and elucidate complex forms of structural variations. We also newly identified SVAs affecting the *APC* gene in two patients with familial adenomatous polyposis, as well as their sites of origin. In addition, we demonstrated that off-target reads from adaptive sampling, which are typically discarded, can be effectively used to accurately genotype common SNPs across the entire genome, enabling the calculation of a polygenic risk score. Furthermore, we identified allele-specific *MLH1* promoter hypermethylation in a Lynch syndrome patient. In summary, our workflow with TAS-LRS can simultaneously capture monogenic risk variants including complex structural variations, polygenic background as well as epigenetic alterations, and will be an efficient platform for genetic disease research and diagnosis.

## Introduction

The advances in sequencing technology have improved the rates of diagnosis of genetic diseases. However, even with whole genome sequencing analysis, the diagnosis rate is still less than half^1^. There has been a lot of interest in long-read sequencing technologies in recent years due to their ability to solve some of the issues related to short-read technologies such as ambiguous alignments on repeat regions and improving SV detection performance^2–4^. Furthermore, long-read sequencing technology also provides DNA modification information, such as 5-methylcytosine and 5-hydroxymethylcytosine, by the signal generated during the sequence of naïve DNA molecules without additional manipulation^5^. Long-read sequencing was, however, plagued by both high error rates and high costs. In order to alleviate these problems, a variety of targeted long-read sequencing has been developed. Examples of these include PCR enrichment^6,7^ and Cas9 target cleavage^8–10^. However, it requires a substantial amount of time during sample preparation.

Recent Oxford Nanopore Technology (ONT) sequencing instruments are equipped with adaptive sampling functions, which adopt or reject DNA molecules based on real-time alignment to sequences in a target region^11–13^. One major advantage of target adaptive sampling long read sequence (TAS-LRS) is that the user only needs to specify the coordinates of the target area, and no prior sample prep is required. The performance of adaptive sampling has been demonstrated in various applications such as Mendelian variant detection, elucidation of complex chromosomal rearrangements, and rapid neurooncology diagnostics^14–17^. Still, the development of a workflow that can systematically identify clinically significant variants is still in its infancy.

We have developed a workflow designed for TAS-LRS sequencing analysis (Figure 1). Our pipeline can precisely detect SNVs and structural variations (SVs) including mobile element insertions such as LINE1, Alu, and SVA (SINE-R/VNTR/Alu). In addition, we have also automated the subsequent detection of pathogenic mutations, making it possible to list candidate variants immediately. In addition, our workflow offers an allele-specific methylation analysis.

**Figure 1:**
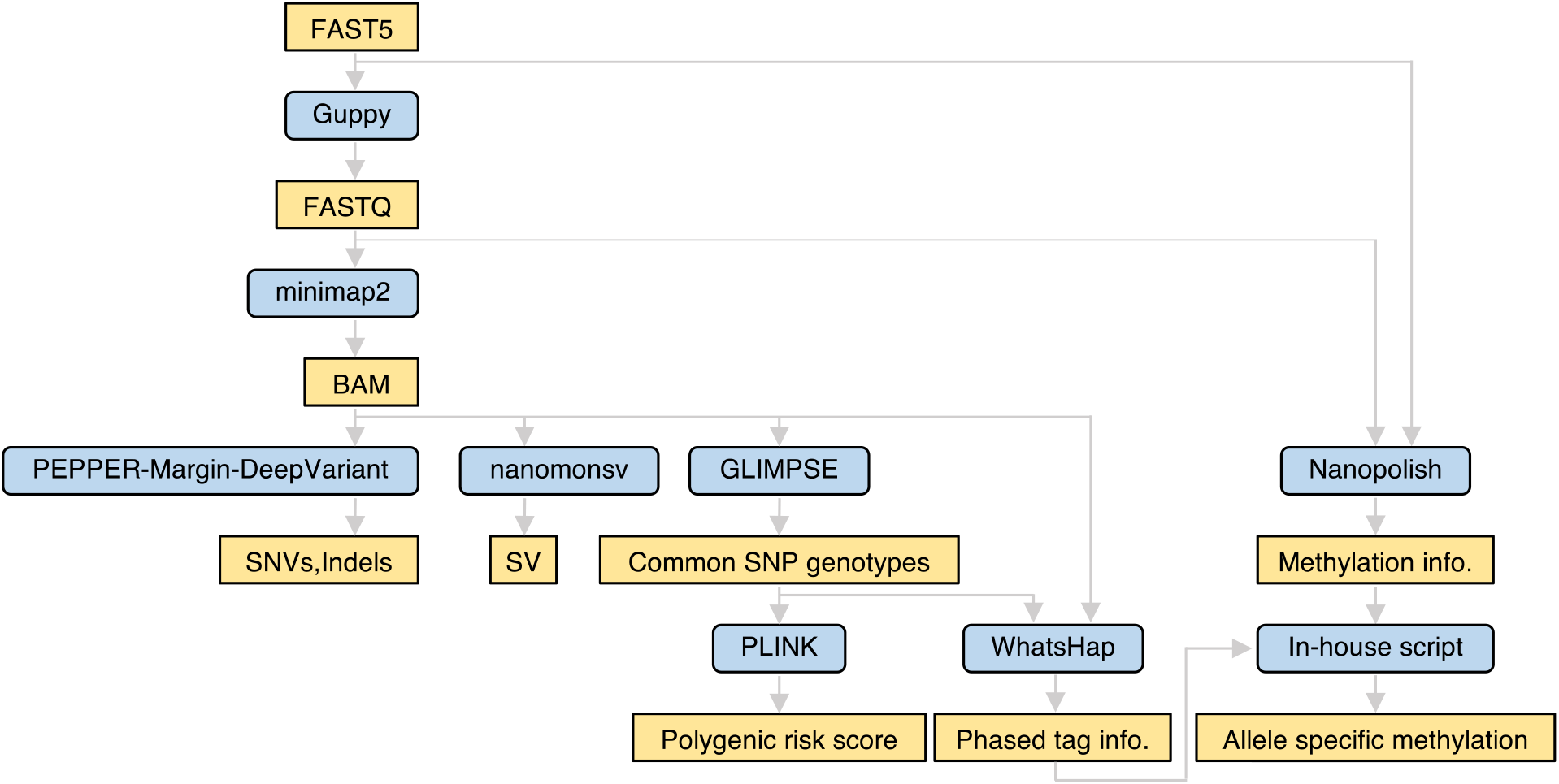
Overview of the workflow for adaptive sampling using nanopore sequencing.

Moreover, we demonstrated that fairly accurate genome-wide common SNP genotyping is possible by making the most of off-target data from TAS-LRS. Recently, various analyzes have shown that not only monogenic pathogenic variants but also polygenic architecture, which is exemplified by Polygenic Risk Score (PRS), have a significant impact on many diseases^18–21^. The calculation of PRS has been typically performed from SNP array or high-coverage whole genome sequencing data. In addition, methods such as low-pass whole genome sequencing with an average sequencing depth of around 1.0x and genotyping from off-target reads in panel sequencing have recently been proposed as a cost-effective option^22–24^. Since adaptive sampling TAS-LRS produces low-coverage sequence data at off-target regions throughout the genome, it would be a reasonable attempt to calculate PRS using them.

We applied our workflow to the TAS-LRS sequencing data from clinically suspected hereditary cancer syndrome patients with the goal of identifying causative variants not found by existing clinical tests, such as panel sequencing, and elucidating the detailed structure of partially detected SVs. Furthermore, we performed the evaluations on SNVs/Indels calling and common SNP genotyping through the comparison with the whole genome sequencing data from short-read sequencing (WG-SRS) data as a benchmark.

## Results

### Selection of 33 patients with suspected hereditary cancer syndromes

In this study, we focused on 33 patients with clinically suspected hereditary cancer syndromes, consisting of 11 familial adenomatous polyposis (FAP), 4 hereditary breast and ovarian cancer (HBOC), 4 retinoblastoma (RB), 4 Li-Fraumeni syndrome (LFS), 2 Lynch syndrome (LS), and 8 other syndromes (Table 1). Two FAP patients had signals of SVs in the *APC* gene from prior analyses (short-read target sequencing (NCC Oncopanel Test^25,26^), or Multiplex Ligation-dependent Probe Amplification (MLPA)), and were analyzed by TAS-LRS to elucidate the detailed form of the SVs precisely. In the two patients with RB, the presence of SVs involving the *RB1* gene was known in advance by FISH and other methods, and TAS-LRS analysis was performed to identify the location of the breakpoints accurately. For 22 patients, high-coverage (range: 26.3× ∼ 31.7×) whole genome sequencing data by Novaseq 6000 platform were available.

**Table 1:** The cohort of patients who participated in this study. WGS: Whole genome sequencing, WTS: Whole transcriptome sequencing, FISH: Fluorescence in situ hybridization, MLPA: Multiplex Ligation dependent Probe Amplification, FAP: Familial adenomatous polyposis, FPC: Familial pancreatic cancer, HAML: Hepatic angiomyolipoma, HBOC: Hereditary breast and ovarian cancer, HDGC: Hereditary diffuse gastric cancer, ICC: intra-hepatic cholangiocarcinoma, LFS: Li-Fraumeni syndrome, LS: Lynch syndrome, MEN1: Multiple endocrine neoplasia type 1, MEN2: Multiple endocrine neoplasia type 2, PHTS: PTEN hamartoma tumor syndrome, PJS: Peutz-Jeghers syndrome, RB: Retinoblastoma.

| Sample ID | Clinical diagnosis | WGS | WTS | NCC Oncopanel Test | G-banding or FISH | MLPA or Sanger | Previous result of genetic analysis |
| --- | --- | --- | --- | --- | --- | --- | --- |
| S1 | LS | ○ | - | ○ | - | - | No pathogenic variants detected. |
| S2 | FAP | ○ | ○ | ○ | - | ○ | A Reciprocal translocation with a breakpoint in *APC* gene was detected by WGS. |
| S3 | RB | ○ | ○ | ○ | ○ | ○ | A Reciprocal translocation with a breakpoint in *RB1* gene was detected by FISH and WGS. |
| S4 | FPC | ○ | - | ○ | - | ○ | No pathogenic variants detected. |
| S5 | FAP | ○ | ○ | ○ | - | ○ | No pathogenic variants detected. |
| S6 | FAP | ○ | ○ | ○ | - | ○ | No pathogenic variants detected. |
| S8 | FAP | ○ | ○ | ○ | - | ○ | No pathogenic variants detected. |
| S9 | MEN1 | ○ | ○ | ○ | - | ○ | No pathogenic variants detected. |
| S11 | MEN2 | ○ | ○ | ○ | - | ○ | No pathogenic variants detected. |
| S12 | LFS | ○ | ○ | ○ | - | ○ | No pathogenic variants detected. |
| S14 | LFS | ○ | - | ○ | - | ○ | No pathogenic variants detected. |
| S15 | HBOC | ○ | - | ○ | - | - | No pathogenic variants detected. |
| S16 | FAP | ○ | ○ | ○ | - | ○ | No pathogenic variants detected. |
| S19 | HAML | ○ | ○ | ○ | - | ○ | No pathogenic variants detected. |
| S20 | FAP | ○ | ○ | ○ | - | ○ | No pathogenic variants detected. |
| S21 | PHTS | ○ | - | ○ | - | ○ | No pathogenic variants detected. |
| S22 | HBOC | ○ | ○ | ○ | - | ○ | No pathogenic variants detected. |
| S23 | RB | ○ | - | ○ | ○ | ○ | No pathogenic variants detected. |
| S24 | RB | ○ | ○ | ○ | ○ | ○ | No pathogenic variants detected. |
| S25 | FPC | ○ | ○ | ○ | - | - | No pathogenic variants detected. |
| S26 | FAP | ○ | ○ | ○ | - | ○ | No pathogenic variants detected. |
| S27 | LFS | ○ | ○ | ○ | - | ○ | No pathogenic variants detected. |
| S28 | PJS | - | - | ○ | - | ○ | No pathogenic variants detected. |
| S29 | ICC | - | - | ○ | - | - | No pathogenic variants detected. |
| S30 | HBOC | - | - | ○ | - | ○ | No pathogenic variants detected. |
| S31 | LFS | - | - | ○ | - | ○ | No pathogenic variants detected. |
| S32 | FAP | - | - | ○ | - | ○ | No pathogenic variants detected. |
| S33 | LS | - | - | ○ | - | ○ | No pathogenic variants detected. |
| S34 | HBOC | - | - | ○ | - | ○ | No pathogenic variants detected. |
| S35 | HDGC | - | - | ○ | - | ○ | No pathogenic variants detected. |
| S36 | FAP | - | - | ○ | - | ○ | No pathogenic variants detected. |
| S37 | FAP | - | - | ○ | - | ○ | A complex structural variation involving *APC* gene was suspected by MLPA. |
| S38 | RB | - | - | ○ | ○ | ○ | A deletion of the *RB1* gene was suspected by MLPA. |

### Summary of sequencing statistics

The median depth of on-target and off-target regions were 23.0 (range: 5.0-44.1) and 2.3 (range: 0.66-6.3), respectively. The median enrichment of the on-target regions compared to the off-target regions was 10.3 (range: 5.6-14.7) (Figure 2a). 17 out of 33 samples had 20x or greater depth of coverage over 50% of the target regions (Supplementary Figure 1). Two samples had an average depth of coverage of less than 10×, compromising the accuracy of subsequent analyses, including mutation detection. The read N50 of on-target and off-target regions were 8,788 (range: 5,330-11,885) and 587 (range: 475-635) (Figure 2b), respectively.

**Figure 2:**
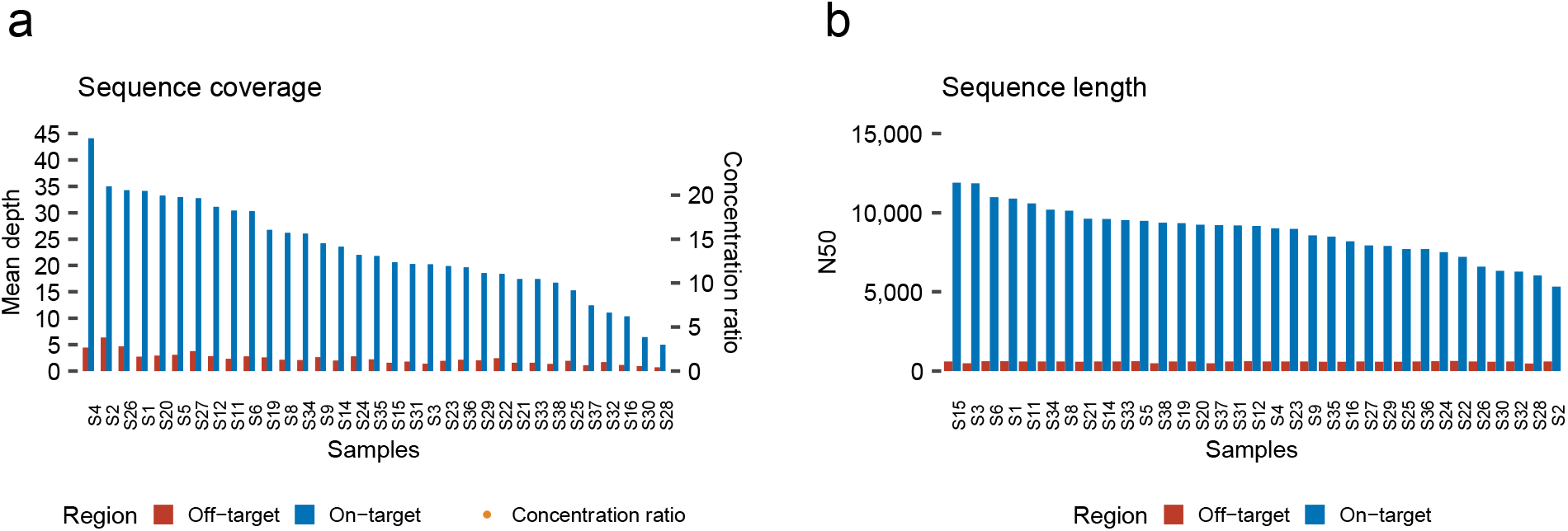
Quality check summary of TAS-LRS sequencing data. (a) Sequence coverage of on-target and off-target regions and concentration ratio (ratio of on-target to off-target coverage) for each sample. Samples were ordered by on-target coverage. (b) N50 statistics calculated for on-target and off-target region for each sample. Samples were ordered by on-target N50 values.

### Evaluation of SNVs/Indels detection

A median of 15,398 (range: 8,633-16,992) SNVs/Indels were detected in the target regions in TAS-LRS (Supplementary Data 2). To evaluate the accuracy of the detection of SNVs/Indels, we compared the detection between TAS-LRS and high-coverage whole-genome short-read sequence (WG-SRS) for 22 cases where matched WG-SRS data were available. For SNVs, the results of the TAS-LRS and WG-SRS were fairly consistent. Setting the SNVs identified from the WG-SRS platform as golden datasets, the median recall and precision for TAS-LRS were 98.8% (range: 90.0-99.4%) and 98.2% (range: 94.7-98.5%), respectively (Figure 3a). The recall of SNVs decreased as the depth of coverage decreased (Supplementary Figure 2), as demonstrated by one case with very low sequence coverage. On the other hand, regarding Indels, the results for the TAS-LRS and WG-SRS were largely different, probably because slippage errors are much more abundant in Oxford Nanopore Technology compared to the Illumina-based short-read platform (Figure 3b).

**Figure 3:**
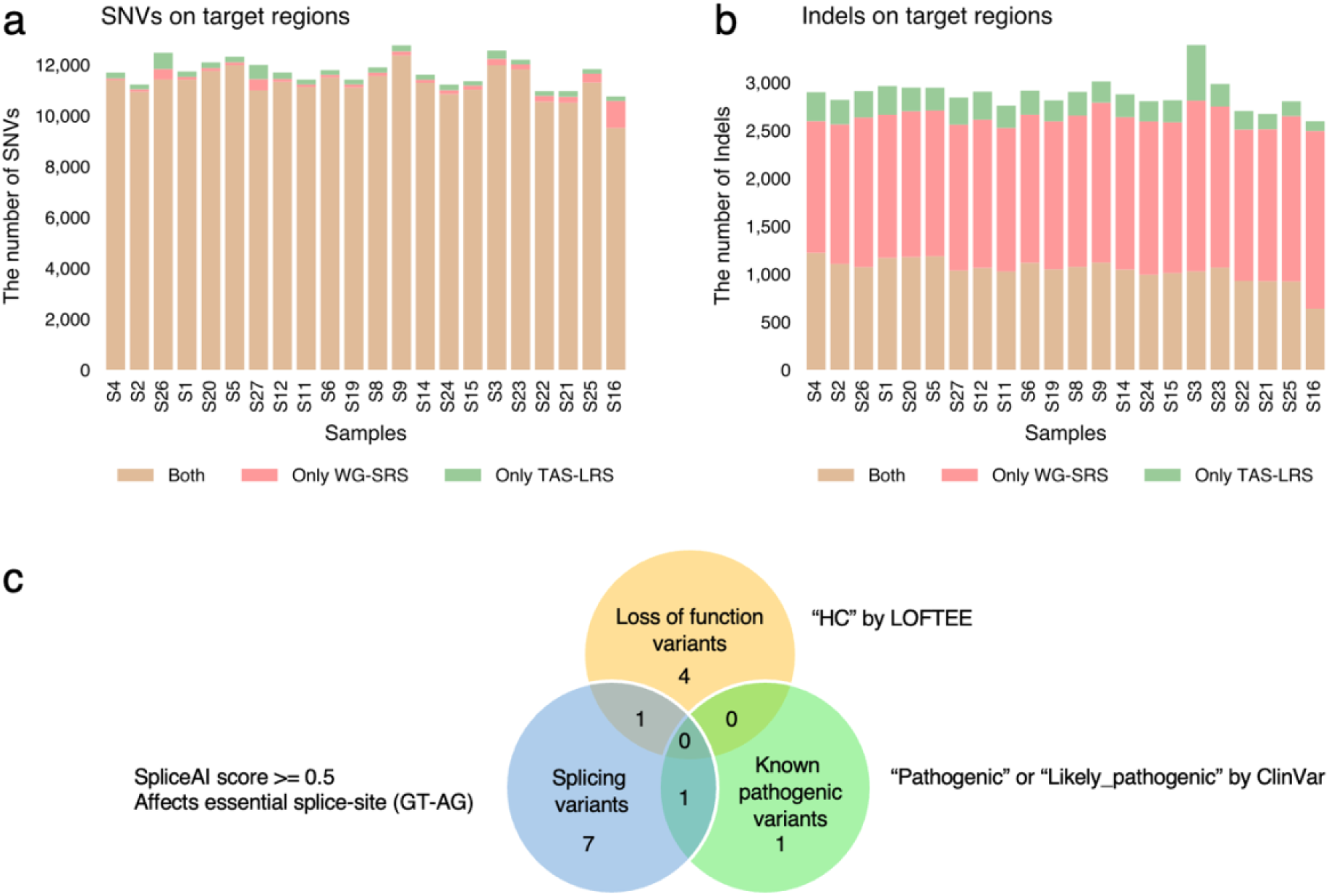
Summary of SNVs/Indels detected in the target region. (a, b) The number of SNVs (a) and indels (b) for each sample stratified by whether they were detected by TAS-LRS, WG-SRS, or both. (c) Venn diagram showing the categories of putative pathogenic variants identified (known pathogenic, loss-of-function, and splicing variants). See also Supplementary Figure 2.

### Extraction of candidate pathogenic SNVs/Indels

From the list of SNVs/Indels detected in the target regions, we extracted candidate pathogenic variants by combining various information such as population allele frequencies, registries of known relationships among variants and diseases (ClinVar^27^), and functional prediction tools (see Methods for detail), and 14 putative pathogenic variants were identified (Figure 3c, Supplementary Figure 3). Two variants were known pathogenic variants registered in ClinVar. Five were high confidence loss of function variants according to LOFTEE^28^. Nine were predicted to cause aberrant splicing. There were two variants belonging to two categories (Supplementary Data 2). Although these putative pathogenic variants were generally detected by previously performed analyses (NCC Oncopanel Test and WG-SRS), we would like to highlight some potential novel findings below.

In a clinically suspected MEN2 (Multiple Endocrine Neoplasia type 2) case (S11), we identified a potential splicing variant in the *EPCAM* gene (c.556-14A>G), which has been registered as “pathogenic” with two stars in the ClinVar. Furthermore, we identified polymorphisms G691S/S904S in the *RET* gene, whose modifier effects on MEN2 have been investigated in several previous studies^29,30^. Therefore, a combination of heterogeneous effects of various mutations might produce symptoms in this patient.

In a patient with clinically suspected Hereditary Diffuse Gastric Cancer (S35), we detected a G258E missense variant in the *MUTYH* gene, which is annotated as “Pathogenic/Likely pathogenic” with two stars in ClinVar. Although impaired glycosylase activity was demonstrated by functional assay for this variant^31^, *MUTYH* is generally considered to be autosomal recessive, and the other mutation has not yet been detected. The association between this mutation and the disease needs to be further investigated.

We also identified a missense mutation in A189V in the *TP53* gene in a clinically suspected LFS patient (S27), which is registered in ClinVar as “Conflicting interpretations of pathogenicity.” This variant showed relatively high minor allele frequency in East Asian cohorts (1.77×10^−3^ in ToMMo 38KJPN database^32^, and 5.46×10^−4^ in Korea 1K) compared to worldwide cohorts (6.57×10^−6^ in gnomAD v3.1.2^28^). The odds ratios for this variant were modest (1.7 ∼ 1.8) in the previous Japanese breast and colorectal cancer cohort study^33,34^. Therefore, the A189V variant in *TP53*, if any, would have only low penetrance pathogenicity.

### Overview of potentially functional SV detection

Application of nanomonsv and unreliable SV removing filters (see Methods for detail) yielded a total of 80 SVs. Furthermore, subsequent putative pathogenic SV extraction (in short, extracting SVs disrupting coding sequences, see Methods for detail) nominated 12 SV breakpoint junctions, two of which were identified as single breakend SV and were in the intron region of the *APC* gene. In fact, all of them involved *RB1* in two RB patients (S3 and S38) or *APC* in two FAP patients (S2 and S37) (Supplementary Figure 4, 5). In the following, we describe a novel clarification provided by TAS-LRS compared to previous tests.

Our analysis resolved a complex intrachromosomal balanced translocation affecting the *RB1* gene and the *LRMDA* gene and identified precise breakpoints, consistent with the one identified by WG-SRS analysis (Fig. 4a). S38 had been inferred to have deletions of exons 21 to 27 (last exon) via MLPA. We showed a large deletion of 44,362 bp extending from the 20th intron of the *RB1* gene to the adjacent *RCBTB2* gene with exact breakpoint coordinates (Figure 4b).

**Figure 4:**
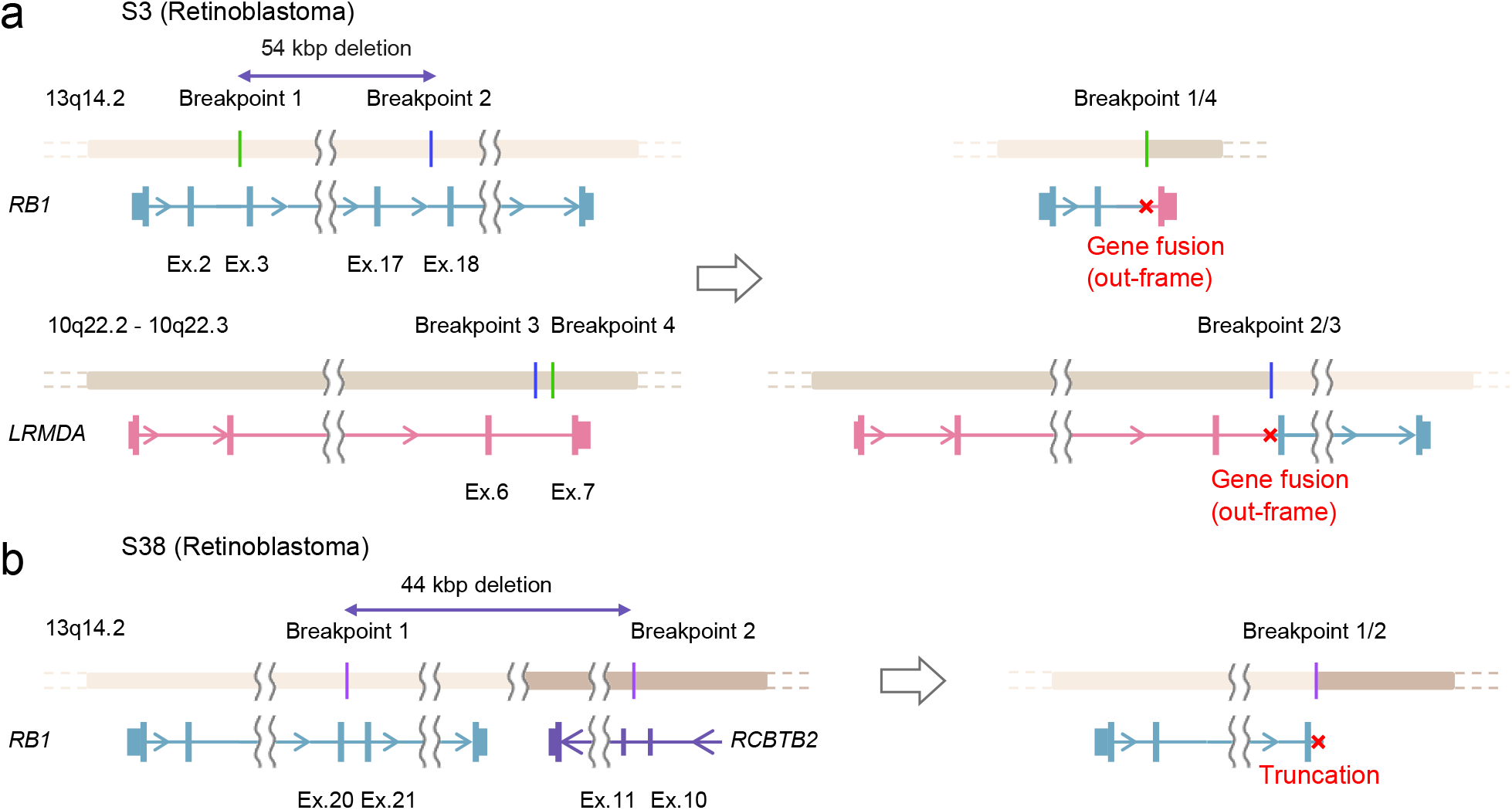
Schematic representation of structural variations of the *RB1* gene in two patients with Retinoblastoma. (a) A balanced translocation involving *RB1* detected in S3 consists of two interchromosomal junctions. One junction connects breakpoint 1 (in the 2nd intron of the *RB1* gene) and breakpoint 4 (in the 6th intron of the *LRMDA* gene), and the other junction juxtaposes breakpoint 3 (in the 17th intron of the *RB1* gene) and breakpoint 4 (in the 6th intron of the *LRMDA* gene). Approximately 54 kbp region between breakpoint 1 and breakpoint 2 in the *RB1* gene was deleted. (b) A deletion spanning a 44 kbp region spanning from the 20th intron of the *RB1* gene to the 10th intron of the *RCBTB2* gene.

Multi-gene panel testing and MLPA had shown partially identified signals indicating SVs on *APC* in S37. However, the overall structure had not yet been elucidated. Our analysis based on TAS-LRS detected two SV breakpoint junctions constituting reciprocal inversions accompanied by 130 kbp deletion. In the other patient with FAP (S2), a reciprocal translocation involving a breakpoint in the *APC* gene was identified (Supplementary Figure 6).

### Two SVA insertions affecting *APC* in FAP patients

We further searched the list of 80 SVs in the phase immediately following the unreliable SV removing filter, focusing in particular on those affecting genes that are well known to be associated with the predicted diseases. This search found a 2,731 bp insertion, which matched to SINE-R/VNTR/Alu (SVA) by RepeatMasker^35^, in the 9th intron of the *APC* genes in a patient of strongly suspected FAP (Figure 5a). SVA is a class of recent mobile elements found only in primates. Mobile element insertions including SVA can cause disease typically by inactivating gene function through abnormal splicing^36,37^. Previous studies have found a number of diseases derived from SVA insertions. Although LINE1, another class of mobile element insertion, has been identified in familial adenomatous polyposis^38–40^, there has been no study that finds SVA insertion in FAP patients as far as we know. The whole transcriptome sequence performed on the same patient revealed significant and specific intron retention at the near exon-intron boundary, implicating the pathogenicity of the SVA insertion in this patient (Figure 5b).

**Figure 5:**
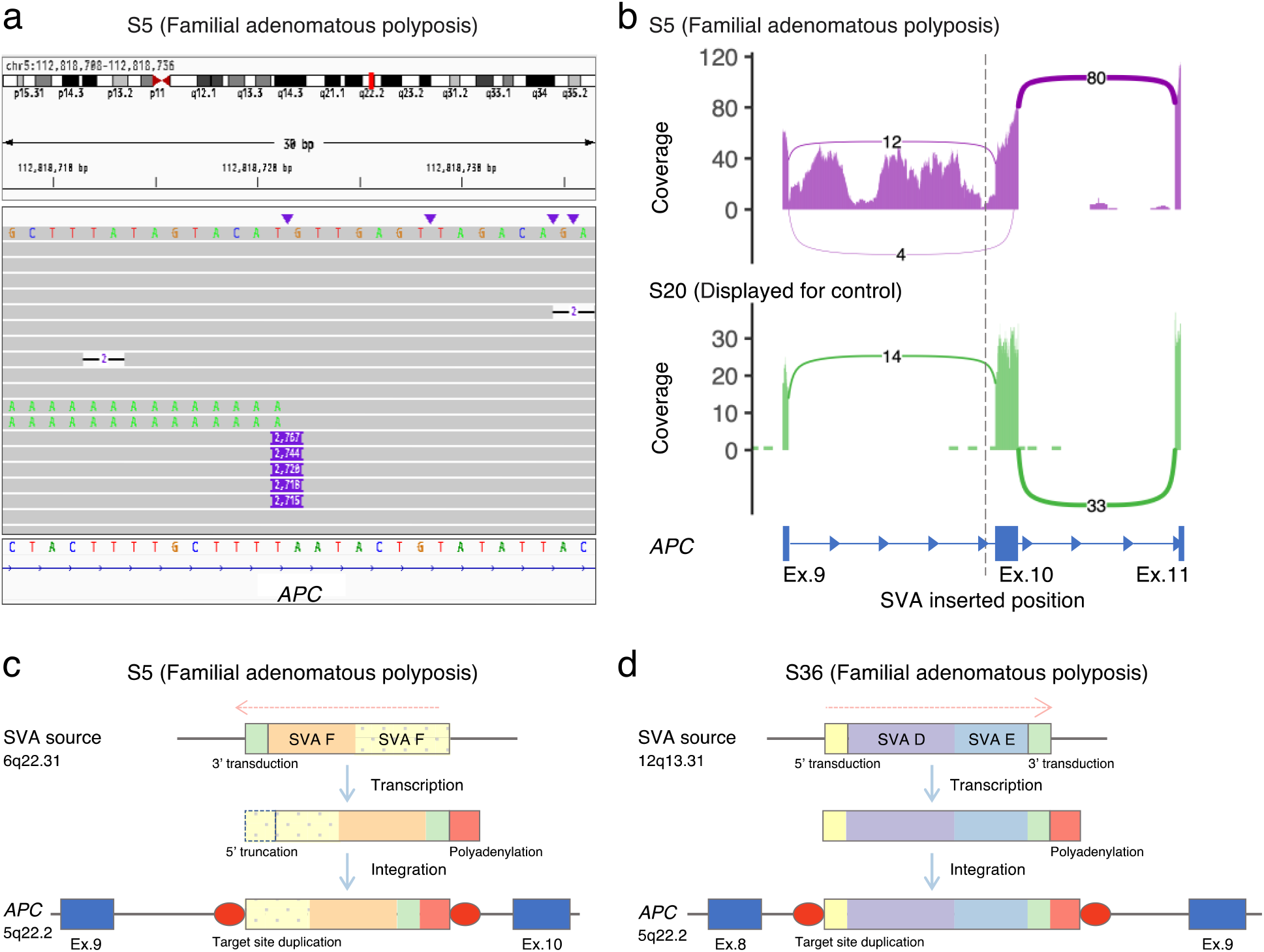
Details of SVA-derived insertion into the intronic region of the *APC* gene in two patients with familial adenomatous polyposis. (a) The IGV displayed long-read sequencing data and transcript sequencing data showing an SVA-derived insertion of 2731 bp in the 9th intron of the *APC* gene. (b) The whole transcriptome sequence showed specific intron retention at the near exon-intron boundary. (c) An SVA inserted into the 9th intron of the *APC* gene in the patient S5, derived from two concatenated human-specific subfamily SVA_F elements located at 6a22.31, which undergoes 5’ truncation and poly(A) tail addition prior to insertion. (d) An SVA inserted into the 8th intron of the *APC* gene in the patient S36, derived from concatenated SVA_D and SVA_E

Next, we investigated the source site by alignment of the SVA sequence to the reference genome using BLAT^41^. The SVA strongly matched the sequence of a region on chromosome 6 (chr6:122,847,699-122,850,317), consisting of two adjacent SVA_F sequences (chr6:122,849,195-122,850,317, chr6:122,847,781-122,849,162) and 82 bp 3′ transduction (chr6:122,847,699-122,847,780), accompanied by a 24 bp polyadenylation tail and 14 bp target site duplication (Figure 5c). The 3′ transduction also contained the predicted conserved polyadenylation signal AATAAA. Since the 5′ end of SVA_F ((CCCTCT)_n_ repeats that is necessary for retrotransposition^42,43^) is truncated, the inserted SVA sequence into *APC* is thought to already lost its trans-mobility capability.

Motivated by this finding, we further investigated for other mobile element insertions by manually investigating the BAM files and identified another 2,678 bp SVA insertion in the 8th intron of the *APC* gene in another FAP patient (Supplementary Figure 7). This inserted sequence was inferred to be derived from a region of chromosome 12 (chr12:8,624,237-8,626,878), which consists of SVA_D (chr12:8,624,321-8,625,515), SVA_E (chr12: 8,625,529-8,626,786), and 91 bp of the 3′ transduction (which included polyadenylation signal), followed by 55 bp polyadenylation tail and 15 bp target site duplication (Figure 5d).

### Verification of genome-wide common SNP genotyping from sequence data from off-target regions

The TAS-LRS provides low-coverage sequencing data even in the off-target regions. There have been several attempts to genotype SNPs genome-wide using off-target sequence data. However, these studies were mostly performed using short-read platforms with few sequence errors, and few attempts have been made on error-prone long reads. Here, we attempted genotyping of common SNPs across the genome (mostly off-target regions), using data from long-read while making the most of off-target reads that would otherwise be discarded.

We performed genome-wide common SNP genotyping on TAS-LRS using GLIMPSE^24^ with a reference panel consisting of 8,570 Japanese genomes from the National Center Biobank Network (NCBN)^44^ project as well as 3,202 genomes from 1000 genomes. The total number of SNPs in the panel was 39,201,938. The number of SNPs genotyped by GLIMPSE for each patient was a median of 1779348.5 (range:1,584,123-1,833,921). The median concordance with WG-SRS was 99.5% (range: 88.4%-99.6%) (Figure 6a, Supplementary Figure 8), showing that common SNP genotyping utilizing low-coverage sequencing reads in the off-target region is fairly accurate in most cases.

**Figure 6:**
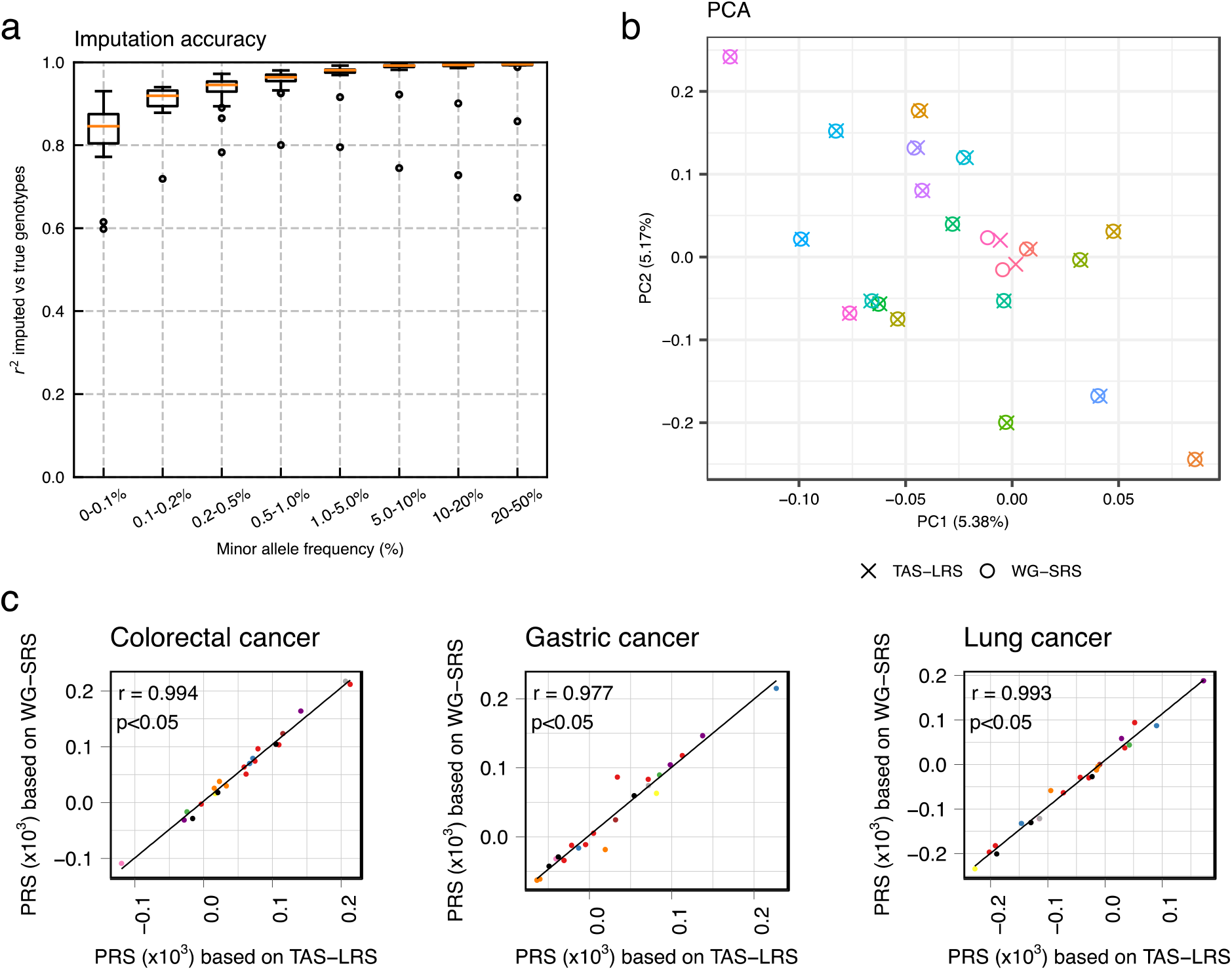
Comparison of genome-wide common SNP genotyping by TAS-LRS (imputation of low-coverage off-target sequencing data using GLIMPSE) compared to WG-SRS (direct variant calling on high-coverage whole-genome sequencing data by GATK). (a) Imputation accuracy of TAS-LRS measured on the chromosome 1 for each minor allele frequency range. Genotyping by WG-SRS was used as the golden standard. See also Supplementary Figure 7. (b) PCA of genotype results from both TAS-LRS and WG-SRS for each individual (distinguished by color). Pairs of the same individuals are clearly clustered, indicating that the batch effect of the difference between the TAS-LRS and WG-SRS platforms has effectively disappeared. One outlier sample that could have originated from different ancestries was excluded. See also Supplementary Figure 8. (c) Comparison of PRSs for three cancers calculated from the genotype by TAS-LRS (X-axis) and WG-SRS (Y-axis). Each point indicates each sample and each color indicates each syndrome name (red: Familial adenomatous polyposis, blue: Familial pancreatic cancer, green: Hepatic angiomyolipoma, purple: Hereditary breast and ovarian cancer, orange: Li-Fraumeni syndrome, yellow: Lynch syndrome, brown: Multiple endocrine neoplasia type 1, pink: Multiple endocrine neoplasia type 2, gray: PTEN hamartoma tumor syndrome, black: Retinoblastoma).

We evaluated the performance of downstream analyses using genotyping results obtained from the above procedure. The result of the principal component analysis (PCA) for the genotype data from both TAS-LRS and WG-SRS showed that identical individuals were strongly clustered, indicating that the batch effect from the platform difference was effectively removed (Figure 6b, Supplementary Figure 9). Next, we calculated a polygenic risk score (PRS) using GWAS summary statistics data^45^ related to various cancer types in the Japanese population and examined the correlation between the scores from TAS-LRS and WG-SRS. PRS calculated by the two platforms showed a strong correlation. The median Pearson correlation for the 12 carcinomas was 0.993 (range: 0.977-0.996) (Figure 6c, Supplementary Figure 10).

These results indicate that genome-wide common SNP genotyping using discarded off-target reads from TAS-LRS is sufficiently accurate for many downstream analyses, such as PRS calculation.

### *MLH1* epimutation in a patient of Lynch syndrome identified via allele-specific methylation analysis

An important advantage of the Oxford Nanopore Technology sequencing data is that epigenetics modifications such as methylation can be obtained for each sequence read and position. Furthermore, by combining the genome-wide genotype obtained in the previous section, which also includes phasing information, it is possible to classify each sequence read by haplotype and obtain allele-specific methylation information. We have implemented a workflow that automatically detects allele-specific methylation regions that are predominantly distant from the pseudo-control data, which were constructed by accumulating methylation information for irrelevant gene regions from 6 samples in which the responsible mutation had been identified. Since Nanopolish does not support R10.4 flowcells, we focused on 29 samples sequenced by R9.4.1 flowcells.

Our analyses revealed constitutional *MLH1* epimutation^46,47^ in a previously undetected patient. This patient was positive for a microsatellite instability test using tumor samples and clinically diagnosed with Lynch syndrome, but no germline mutation was found in multi-gene panel testing. As shown in Figure 7, we visualized the allele-specific methylation status of this patient using our workflow. Our workflow revealed that one allele of the *MLH1* exhibited hypermethylation in the promoter region, suggesting that the *MLH1* gene expression is reduced in one allele. Based on the above, the patient was suspected o*f MLH1* epimutation for the first time.

**Figure 7:**
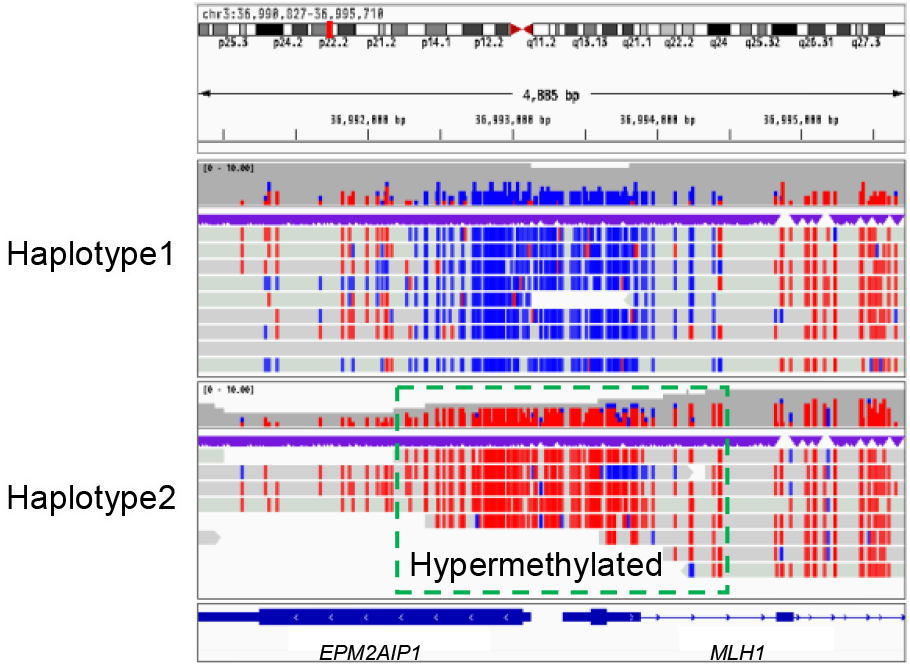
A case of an *MLH1* epimutation in a patient with LS. Alignment view of around the promoter region of the *MLH1* gene. Each read was classified as haplotype 1 or 2 using Whatshap software. The CpG sites of each read are colored red if methylated and blue if not. It can be clearly seen that methylation is increased specifically for haplotype 2.

Also, we detected a patient with FAP, which showed abnormal hypermethylation in the first intron region of the *BARD1* gene. However, their relevance to the disease remains to be investigated (Supplementary Figure 11).

## Discussion

In this study, we demonstrated that TAS-LRS can identify single nucleotide variants with nearly the same accuracy as the short-read platform, and elucidate complex structural variations including mobile element insertions. In addition, we showed that off-target reads can be used to genotype common SNPs genome-wide. Furthermore, it is possible to identify allele-specific methylation aberrations. Thus, a single platform can simultaneously capture the genomic and epigenetic status of monogenic disease-causing genes as well as polygenic effects^48^.

One of the major challenges for LRS at present is the accurate detection of Indels, as in other studies using error-prone long reads. It may require some heuristics such as devising effective post-filtering of Indels which are located in homopolymers. At the same time, measurement technology has continued to make great progress. In fact, the evaluation in this study was mostly performed using the R9.4.1 flowcell, which is a slightly earlier generation, and the accuracy of Indel detection would increase if a newer sequencing kit (Kit V14) were used^49^. In addition, recent ONT duplex technologies, which attach adapters to both strands of a DNA molecule and sequence them from both sides, have been reported to achieve Q30 accuracy. We are optimistic that the problem of LRS Indel detection problem of LRS will be resolved in the near future.

Mobile element insertions such as SVA are notoriously difficult to detect with existing short-read sequencing platforms. In two FAP patients, we successfully identified SVA insertions and their detailed characteristics such as their sites of origin. In fact, the detection of mobile element insertions may contribute to future therapeutic options as well as diagnostics. A groundbreaking study administered a personalized antisense oligonucleotide therapy for a child with Batten disease that targets abnormal splicing caused by a sporadic SVA insertion^50^.

Although a current definitive treatment for FAP patients is prophylactic colectomy before colorectal cancer develops, it has many limitations such as surgical complications and decreased quality of life. Tailor-made medicine such as an antisense oligonucleotide by examining the genomes of FAP patients and other hereditary tumor-related diseases may be an alternative treatment in the future.

We have demonstrated that genotyping of common SNPs can be performed accurately using off-target reads. In this study, we focused on genotyping SNPs with relatively high minor allele frequencies (MAFs). The accuracy of genotyping at rarer variants will continue to improve in the future as large-scale biobank data accumulate and reference panels become more well-stocked, which may even allow for the identification of pathogenic variants with intermediate MAF despite those being located in the off-target region. An increase in concentration ratio due to improvement in sequencing accuracy and real-time alignment computational time may reduce off-target reads for genome-wide genotyping. In such cases, we will be able to explicitly adjust settings, such as specifying a time or the number of pores in which adaptive sampling is performed.

Allele-specific methylation status could be assessed without the need for experiments such as bisulfite conversion. The current study focused on target regions of known cancer predisposition genes. However, as a previous report demonstrated that methylation information from low-coverage sequencing can reveal cancer subtypes^51^, there is a possibility that information from off-target regions can be effectively used in the future.

In this study, we focused on identifying variants in the germline genome. On the other hand, somatic mutations are more difficult to detect than germline mutations due to the impurity of the tumor, the presence of subclones, and the low frequency of variant alleles. However, it is expected that the accuracy of sequencing and the amplification rate of adaptive sampling will increase in the future due to the development of products such as flow cells and reagents and the improvement of various analytical instruments. We are optimistic that reliable somatic mutation detection, including Indels, will eventually be achievable in the near future, and that TAS-LRS will replace the current panel sequence based on a short-read platform.

## Conclusions

We showed that our workflow enables SNVs/Indels detection, methylation status detection, and genome-wide common SNP genotyping. Moreover, our workflow is not just long-read target sequencing to cover the disadvantages of short-read sequencing, but also SNP array sequencing of whole genomic regions and haplotype-by-haplotype methylation analysis of target regions without additional operations. This striking method can be flexibly executed by simply specifying the target regions computationally.

## Methods

### Adaptive sampling sequencing with GridION

We curated 147 cancer predisposition genes from the literature. To generate the BED file specifying a target region, the maximal regions of all the corresponding transcripts registered in the GENCODE Basic gene annotation Release 38 with a margin of 10 kbp for each gene were set. The resulting target region size was 16,122,639 bp (See Supplementary Data 1).

DNA and RNA were obtained after the isolation of plasma from blood samples. For each sample, 10 μg of genomic DNA was sheared using a Covaris g-TUBE by centrifugation at 4,200 rpm for 60 seconds, followed by inversion and centrifugation again at 4,200 rpm for 60 seconds. DNA for sequencing was prepared using the ONT Ligation Kit (SQK-LSK110) according to the manufacturer’s instructions. Each library was loaded mostly on R9.4.1 FLO-MIN106D (29 patients) and rarely on R10.4 FLO-MIN112 (4 patients) flow cells. Sequencing was performed in “hac” mode using the adaptive sampling option for 72 hours with two additional library loadings once every 24 hours after nuclease flushing of a flow cell using the Flow Cell Wash Kit (EXP-WSH004).

### Alignment of the sequencing data

FAST5 files were base-called using the high-accuracy model in Guppy (ver.6.0.7). All FASTQ files were aligned using Minimap2 (ver.2.22-r1101)^52^ with the “-ax map-ont -t 8 -p 0.1 -- MD” option to the GRCh38 human reference genome, and converted into BAM files, and sorted and indexed with Samtools (ver.1.13)^53^.

### Detection, annotation, and prioritization of SNVs/Indels

Denovo SNV, Indels detection were performed using PEPPER-Margin-DeepVariant (ver.0.8.0)^54^, via “run_pepper_margin_deepvariant call_variant” command with the “--ont_r9_guppy5_sup” mode, and filtered out the SNVs/Indels with <= 10 quality values by the software. Then, using Ensembl Variant Effect Predictor (VEP) ver.105.0^55^ and our in-house custom scripts, we annotated SNVs/Indels with the deleteriousness prediction (CADD ver.1.6^56^, ClinVar^27^, and LOFTEE^28^), the splicing prediction (SpliceAI^57^), splicing variant database (SAVNet^58^ and IRAVDB^59^), and the population allele frequencies (gnomAD ver.3.1.2^28^ and ToMMo 14JPN^32^).

To select candidates of pathogenic SNVs/Indels, we first removed those commonly found in general populations (allele frequencies equal to or greater than 0.01 by gnomAD v.3.1.2 and ToMMo 38KJPN)^28,60,61^. Then, variants satisfying either of the following conditions were extracted as putative pathogenic variants.

1. Annotated as “Pathogenic” or “Likely pathogenic” by ClinVar^27^.
2. Affecting essential splice-site (GT-AG) or those whose delta score by SpliceAI^57^ is 0.50 or greater.
3. Loss of function variants and deemed as “High-confidence (HC)” by LOFTEE^28^.

### Detection, annotation, and prioritization of SVs

SVs were detected using nanomonsv (ver.0.5.0b2)^62^. After “nanomonsv parse” was performed for each BAM file, we executed “nanomonsv get” with “--single_bnd --use_racon” option to identify single breakend SVs. We also included a dummy matched control and the control panel data created from the sequencing data from Human Pangenome Reference Consortium (provided by Zenodo (https://zenodo.org/record/7017953#.Y8uP-uxByZw). For the dummy match control, we used whole-genome long-read sequencing data (by Oxford Nanopore Technology, base-called with Guppy 5.0.11) of a Japanese male (NA18989) provided by the Human Genome Structural Variation Consortium^63,64^. Then, we filtered out SVs that satisfy the following conditions (unreliable SV removing filter):

1. One or both of the SV breakpoints are present in the unplaced contig, decoy sequence.
2. Neither SV breakpoint is located within the target region.
3. Deletion, insertion, and tandem duplication type SVs included in the simple repeat region (including 10 bp margin).

Furthermore, the following categories of SVs were extracted as putative pathogenic SVs (putative pathogenic SVs extraction):

1. Deletions involving the coding regions of the 147 target genes
2. Duplications that alter the coding sequences of 147 genes. More specifically, those that have at least one breakpoint within the target genes excluding untranslated regions and the amplified regions span the coding sequences.
3. Inversions or translocations that disrupt the coding sequences of 147 genes. That is, those that have at least one of the breakpoints in a gene region excluding the untranslated regions.

We also included single breakend SVs detected by nanomonsv with the “--single_bnd -- use_racon” options.

### Common SNP genotype calling using GLIMPSE

First, we generated a reference panel using 8,570 short-read platform whole genome sequencing data collected from National Center Biobank Network (NCBN)^44^ as well as 3,202 sequencing data from1000 Genomes Project. After alignment, and variant calling using a standard pipeline, we retained variants meeting genotypic HWE (Hardy-Weinberg Equilibrium) goodness-of-fit test P-value ≥ 0.0001.

Then, we performed genotyping for each BAM file from TAS-LRS using GLIMPSE (ver.1.1.1)^24^ using the above reference panel. First, multiallelic sites of VCF files were split into biallelic records using the “bcftools norm -m -any’’ command, then converted to tsv files using “bcftools query -f’%CHROM\t%POS \t%REF,%ALT\n’ ${REFVCF} | bgzip -c” command and indexed the tsv file using “tabix -s1 -b2 -e2” command. In addition to the options described in the tutorial, genotype likelihoods for a single individual at specific positions were computed using bcftools mpileup with the “-X ont” option and bcftools call with the “-P 0.01” option. The GLIMPSE_chunk command to generate imputation regions for each chromosome was performed with the “--window-size 2000000” and “--buffer-size 200000” options. The GLIMPSE_phase command to impute and phase a whole chromosome was run using default parameters. Fine-scale genetic maps were downloaded from the URL https://zenodo.org/record/4078748. The GLIMPSE_ligate and the GLIMPE_sample commands were performed as default parameters.

To compare genotyping results between TAS-LRS using GLIMPSE and WG-SRS using GATK, we excluded simple repeat regions, segmental duplication regions, and regions to which alternative haplotype sequences match, which we downloaded from the annotation database for the UCSC Genome Browser (https://hgdownload.soe.ucsc.edu/goldenPath/hg38/database). We used GLIMPSE2_concordance^65^ tool to evaluate the accuracy of genotyping. We followed the GLIMPSE2 tutorial (https://odelaneau.github.io/GLIMPSE/docs/tutorials/getting_started/) and checked the imputation accuracy using the “GLIMPSE2_concordance” command with the “--min-val-dp 8 --min-val-gl 0.9999 --bins 0.000 0.001 0.002 0.005 0.010 0.050 0.100 0.200 0.500” options.

### Allele-specific methylation analysis

First, we classified each read in the BAM file using WhatsHap (ver.1.4)^66^ (“whatshap haplotag” command was used to tag reads by haplotype with “--skip-missing-contigs” and “--ignore-read-groups” options) based on the phased common SNP genotype VCF file obtained using GLIMPSE in the previous subsection. Nanopolish (ver.0.13.3)^67^ was then used for the methylation calling. After indexing the FASTA file, the command “nanopolish call-methylation” was executed. Next, the output file containing the log-likelihood ratio of methylation per read and CG dinucleotide was divided into three files (“haplotype 1”, “haplotype 2”, and “haplotype unclassified”). Finally, the allele-specific methylation frequencies for each CG dinucleotide were calculated using calculate_methylation_frequency.py (downloaded from the Nanopolish GitHub website) on the above split files with the ‘-s’ option.

To convert the BAM file for methylation visualization via Integrative Genomic Viewer^68^, we used the convert_bam_for_methylation.py script (https://github.com/timplab/nanopore-methylation-utilities)^69^.

### The comparison of polygenic risk score

PRS was calculated using PLINK (ver. 1.90 beta)^70^ according to the tutorial page (https://choishingwan.github.io/PRS-Tutorial/plink/)^71^. We used GWAS summary statistics^45^ for 12 cancer types (breast cancer, biliary tract cancer, cervical cancer, colorectal cancer, endometrial cancer, esophageal cancer, gastric cancer, hepatocellular cancer, lung cancer, ovarian cancer, pancreatic cancer, and prostate cancer) downloaded from Biobank Japan (http://jenger.riken.jp/result) as base data. We converted the coordinate of these summary statistics to hg38 using the UCSC LiftOver^72^ tool and processed them according to the tutorial. For the target data, we used VCF files for common SNPs by GLIMPSE for TAS-LRS merged across samples. We also used a phased VCF file for the corresponding individuals extracted from the reference panel produced by BEAGLE for WG-SRS for corresponding individuals. For the QC of the target data, we mostly followed the tutorial page. However, we modified the option of -- indep-pairwise to 50000 5000 0.50 when removing highly correlated SNPs. Also, we did not perform sample exclusion by the standard deviation of the F coefficient due to the small sample size. Finally, we performed clumping and calculated PRS for each cancer. Then, we compared the PRS calculated by TAS-LRS and WG-SRS for each individual.

### Dimension reduction with principal components analysis

Principal components analysis (PCA) was performed using PLINK (ver. 1.90 beta)^70^. First, the linkage pruning was performed by PLINK with the option of --indep-pairwise 50 10 0.10 --geno 0.01 --maf 0.1 –freq.” Then, we next performed PLINK with “--pca” to obtain eigen vector and values.

## Data Availability

The workflow used in this study is available on GitHub at https://github.com/ncc-gap/ASWorkflow. The raw Oxford Nanopore sequence data via target adaptive sampling used in this study will be available through the public sequence repository service.

## Acknowledgments

We thank all patients who provided samples for this study.

## Author’s Contributions

YS and MH designed the study. WN and YS developed the workflow for sequencing analysis with the help of NI, KC, AO, RNM and MS. WN, MH, SO, MU, ST, NS, SI, and TY contributed to data collection. WN and YS interpreted the data and result with the assistance of MH, SO, MU, ST, NS, SI, and TY. WN and YS generated figures. YS and WN wrote the manuscript.

## Ethics approval and consent to participate

The research protocol was approved by the Ethics Committee of the National Cancer Center Hospital (Tokyo, Japan) (approval #2013-303). Written informed consent for clinical genetic testing and genomic analysis was obtained from patients. All patient information was deidentified.

## Consent for publication

Not applicable.

## Supplementary Information

### Supplementary Data

**Supplementary Data 1:** List of cancer predisposition genes selected by the National Cancer Center Hospital.

**Supplementary Data 2:** List of SNVs/Indels detected by PEPPER-Margin-DeepVariant remaining after filtering.

**Supplementary Data 3:** List of SVs detected by nanomonsv remaining after putative pathogenic SVs extraction filtering.

**Supplementary Data 4:** List of single breakend SVs detected by nanomonsv (with “--single_bnd option) remaining after putative pathogenic SVs extraction filtering.

**Supplementary Data 5:** List of the members of the NCBN Controls WGS Consortium.

## References

1. 100,000 Genomes Project Pilot Investigators et al. 100,000 Genomes Pilot on Rare-Disease Diagnosis in Health Care - Preliminary Report. N. Engl. J. Med. 385, 1868–1880 (2021).

2. Wang, Y., Zhao, Y., Bollas, A., Wang, Y. & Au, K. F. Nanopore sequencing technology, bioinformatics and applications. Nat. Biotechnol. 39, 1348–1365 (2021).

3. Beyter, D. et al. Long-read sequencing of 3,622 Icelanders provides insight into the role of structural variants in human diseases and other traits. Nat. Genet. 53, 779–786 (2021).

4. Jiang, T. et al. Long-read-based human genomic structural variation detection with cuteSV. Genome Biol. 21, 189 (2020).

5. Logsdon, G. A., Vollger, M. R. & Eichler, E. E. Long-read human genome sequencing and its applications. Nat. Rev. Genet. 21, 597–614 (2020).

6. Karamitros, T. & Magiorkinis, G. A novel method for the multiplexed target enrichment of MinION next generation sequencing libraries using PCR-generated baits. Nucleic Acids Res. 43, e152 (2015).

7. Yamaguchi, K. et al. Application of targeted nanopore sequencing for the screening and determination of structural variants in patients with Lynch syndrome. J. Hum. Genet. 66, 1053–1060 (2021).

8. Gilpatrick, T. et al. Targeted nanopore sequencing with Cas9-guided adapter ligation. Nat. Biotechnol. 38, 433–438 (2020).

9. Gabrieli, T. et al. Selective nanopore sequencing of human BRCA1 by Cas9-assisted targeting of chromosome segments (CATCH). Nucleic Acids Res. 46, e87 (2018).

10. Karamitros, T. & Magiorkinis, G. Multiplexed Targeted Sequencing for Oxford Nanopore MinION: A Detailed Library Preparation Procedure. Methods Mol. Biol. 1712, 43–51 (2018).

11. Loose, M., Malla, S. & Stout, M. Real-time selective sequencing using nanopore technology. Nat. Methods 13, 751–754 (2016).

12. Payne, A. et al. Readfish enables targeted nanopore sequencing of gigabase-sized genomes. Nat. Biotechnol. 39, 442–450 (2021).

13. Kovaka, S., Fan, Y., Ni, B., Timp, W. & Schatz, M. C. Targeted nanopore sequencing by real-time mapping of raw electrical signal with UNCALLED. Nat. Biotechnol. 39, 431–441 (2021).

14. Miller, D. E. et al. Targeted long-read sequencing identifies missing disease-causing variation. Am. J. Hum. Genet. 108, 1436–1449 (2021).

15. Mariya, T. et al. Target enrichment long-read sequencing with adaptive sampling can determine the structure of the small supernumerary marker chromosomes. J. Hum. Genet. 67, 363–368 (2022).

16. Patel, A. et al. Rapid-CNS2: rapid comprehensive adaptive nanopore-sequencing of CNS tumors, a proof-of-concept study. Acta Neuropathol. 143, 609–612 (2022).

17. Yamada, M. et al. Diagnosis of Prader-Willi syndrome and Angelman syndrome by targeted nanopore long-read sequencing. Eur. J. Med. Genet. 66, 104690 (2023).

18. Lewis, C. M. & Vassos, E. Polygenic risk scores: from research tools to clinical instruments. Genome Med. 12, 44 (2020).

19. Torkamani, A., Wineinger, N. E. & Topol, E. J. The personal and clinical utility of polygenic risk scores. Nat. Rev. Genet. 19, 581–590 (2018).

20. Hao, L. et al. Development of a clinical polygenic risk score assay and reporting workflow. Nat. Med. 28, 1006–1013 (2022).

21. Wand, H. et al. Improving reporting standards for polygenic scores in risk prediction studies. Nature 591, 211–219 (2021).

22. Homburger, J. R. et al. Low coverage whole genome sequencing enables accurate assessment of common variants and calculation of genome-wide polygenic scores. Genome Med. 11, 74 (2019).

23. Ho, W.-K. et al. European polygenic risk score for prediction of breast cancer shows similar performance in Asian women. Nat. Commun. 11, 3833 (2020).

24. Rubinacci, S., Ribeiro, D. M., Hofmeister, R. J. & Delaneau, O. Efficient phasing and imputation of low-coverage sequencing data using large reference panels. Nature Genetics vol. 53 120–126 Preprint at https://doi.org/10.1038/s41588-020-00756-0 (2021).

25. Sunami, K. et al. Feasibility and utility of a panel testing for 114 cancer-associated genes in a clinical setting: A hospital-based study. Cancer Sci. 110, 1480–1490 (2019).

26. Kato, M. et al. A computational tool to detect DNA alterations tailored to formalin-fixed paraffin-embedded samples in cancer clinical sequencing. Genome Med. 10, 44 (2018).

27. Landrum, M. J. et al. ClinVar: improving access to variant interpretations and supporting evidence. Nucleic Acids Res. 46, D1062–D1067 (2018).

28. Karczewski, K. J. et al. The mutational constraint spectrum quantified from variation in 141,456 humans. Nature 581, 434–443 (2020).

29. Robledo, M. et al. Polymorphisms G691S/S904S of RET as Genetic Modifiers of MEN 2A1. Cancer Res. 63, 1814–1817 (2003).

30. Gil, L. et al. Genetic analysis of RET, GFR alpha 1 and GDNF genes in Spanish families with multiple endocrine neoplasia type 2A. Int. J. Cancer 99, 299–304 (2002).

31. Yanaru-Fujisawa, R. et al. Genomic and functional analyses of MUTYH in Japanese patients with adenomatous polyposis. Clin. Genet. 73, 545–553 (2008).

32. Tadaka, S. et al. jMorp updates in 2020: large enhancement of multi-omics data resources on the general Japanese population. Nucleic Acids Res. 49, D536–D544 (2021).

33. Fujita, M. et al. Population-based Screening for Hereditary Colorectal Cancer Variants in Japan. Clin. Gastroenterol. Hepatol. 20, 2132–2141.e9 (2022).

34. Momozawa, Y. et al. Germline pathogenic variants of 11 breast cancer genes in 7,051 Japanese patients and 11,241 controls. Nat. Commun. 9, 4083 (2018).

35. A., S. A. F. Repeat-Masker Open-3.0. http://www.repeatmasker.org (2004).

36. Payer, L. M. & Burns, K. H. Transposable elements in human genetic disease. Nat. Rev. Genet. 20, 760–772 (2019).

37. Hancks, D. C. & Kazazian, H. H., Jr. Roles for retrotransposon insertions in human disease. Mob. DNA 7, 9 (2016).

38. Taniguchi-Ikeda, M. et al. Pathogenic exon-trapping by SVA retrotransposon and rescue in Fukuyama muscular dystrophy. Nature 478, 127–131 (2011).

39. Miki, Y. et al. Disruption of the APC gene by a retrotransposal insertion of L1 sequence in a colon cancer. Cancer Res. 52, 643–645 (1992).

40. Scott, E. C. et al. A hot L1 retrotransposon evades somatic repression and initiates human colorectal cancer. Genome Res. 26, 745–755 (2016).

41. James Kent, W. BLAT—The BLAST-Like Alignment Tool. Genome Res. 12, 656–664 (2002).

42. Hancks, D. C., Mandal, P. K., Cheung, L. E. & Kazazian, H. H., Jr. The minimal active human SVA retrotransposon requires only the 5’-hexamer and Alu-like domains. Mol. Cell. Biol. 32, 4718–4726 (2012).

43. Raiz, J. et al. The non-autonomous retrotransposon SVA is trans-mobilized by the human LINE-1 protein machinery. Nucleic Acids Res. 40, 1666–1683 (2012).

44. Kawai, Y. et al. Exploring the genetic diversity of the Japanese Population: Insights from a Large-Scale Whole Genome Sequencing Analysis. bioRxiv 2023.01.23.525133 (2023) doi:10.1101/2023.01.23.525133.

45. Ishigaki, K. et al. Large scale genome-wide association study in a Japanese population identified 45 novel susceptibility loci for 22 diseases. bioRxiv 795948 (2019) doi:10.1101/795948.

46. Ward, R. L., Dobbins, T., Lindor, N. M., Rapkins, R. W. & Hitchins, M. P. Identification of constitutional MLH1 epimutations and promoter variants in colorectal cancer patients from the Colon Cancer Family Registry. Genet. Med. 15, 25–35 (2013).

47. Goodfellow, P. J. et al. Combined Microsatellite Instability, MLH1 Methylation Analysis, and Immunohistochemistry for Lynch Syndrome Screening in Endometrial Cancers From GOG210: An NRG Oncology and Gynecologic Oncology Group Study. J. Clin. Oncol. 33, 4301–4308 (2015).

48. Gusev, A., Groha, S., Taraszka, K., Semenov, Y. R. & Zaitlen, N. Constructing germline research cohorts from the discarded reads of clinical tumor sequences. Genome Med. 13, 179 (2021).

49. Sereika, M. et al. Oxford Nanopore R10. 4 long-read sequencing enables the generation of near-finished bacterial genomes from pure cultures and metagenomes without short-read or reference polishing. Nat. Methods 19, 823–826 (2022).

50. Kim, J. et al. Patient-Customized Oligonucleotide Therapy for a Rare Genetic Disease. N. Engl. J. Med. 381, 1644–1652 (2019).

51. Djirackor, L. et al. Intraoperative DNA methylation classification of brain tumors impacts neurosurgical strategy. Neurooncol Adv 3, vdab149 (2021).

52. Li, H. Minimap2: pairwise alignment for nucleotide sequences. Bioinformatics 34, 3094– 3100 (2018).

53. Danecek, P. et al. Twelve years of SAMtools and BCFtools. Gigascience 10, (2021).

54. Shafin, K. et al. Haplotype-aware variant calling with PEPPER-Margin-DeepVariant enables high accuracy in nanopore long-reads. Nat. Methods 18, 1322–1332 (2021).

55. McLaren, W. et al. The Ensembl Variant Effect Predictor. Genome Biol. 17, 122 (2016).

56. Rentzsch, P., Schubach, M., Shendure, J. & Kircher, M. CADD-Splice—improving genome-wide variant effect prediction using deep learning-derived splice scores. Genome Medicine vol. 13 Preprint at https://doi.org/10.1186/s13073-021-00835-9 (2021).

57. Jaganathan, K. et al. Predicting Splicing from Primary Sequence with Deep Learning. Cell 176, 535–548.e24 (2019).

58. Shiraishi, Y. et al. A comprehensive characterization of cis-acting splicing-associated variants in human cancer. Genome Res. 28, 1111–1125 (2018).

59. Shiraishi, Y. et al. Systematic identification of intron retention associated variants from massive publicly available transcriptome sequencing data. Nat. Commun. 13, 5357 (2022).

60. Yamaguchi-Kabata, Y. et al. Evaluation of reported pathogenic variants and their frequencies in a Japanese population based on a whole-genome reference panel of 2049 individuals. J. Hum. Genet. 63, 213–230 (2018).

61. Nagasaki, M. et al. Rare variant discovery by deep whole-genome sequencing of 1,070 Japanese individuals. Nat. Commun. 6, 8018 (2015).

62. Shiraishi, Y. et al. Precise characterization of somatic structural variations and mobile element insertions from paired long-read sequencing data with nanomonsv. bioRxiv 2020.07.22.214262 (2021) doi:10.1101/2020.07.22.214262.

63. Ebert, P. et al. Haplotype-resolved diverse human genomes and integrated analysis of structural variation. Science 372, (2021).

64. Chaisson, M. J. P. et al. Multi-platform discovery of haplotype-resolved structural variation in human genomes. Nat. Commun. 10, 1784 (2019).

65. Rubinacci, S., Hofmeister, R., da Mota, B. S. & Delaneau, O. Imputation of low-coverage sequencing data from 150,119 UK Biobank genomes. bioRxiv 2022.11.28.518213 (2022) doi:10.1101/2022.11.28.518213.

66. Martin, M. et al. WhatsHap: fast and accurate read-based phasing. bioRxiv 085050 (2016) doi:10.1101/085050.

67. Simpson, J. T. et al. Detecting DNA cytosine methylation using nanopore sequencing. Nat. Methods 14, 407–410 (2017).

68. Thorvaldsdóttir, H., Robinson, J. T. & Mesirov, J. P. Integrative Genomics Viewer (IGV): high-performance genomics data visualization and exploration. Brief. Bioinform. 14, 178– 192 (2013).

69. Lee, I. et al. Simultaneous profiling of chromatin accessibility and methylation on human cell lines with nanopore sequencing. Nat. Methods 17, 1191–1199 (2020).

70. Chang, C. C. et al. Second-generation PLINK: rising to the challenge of larger and richer datasets. GigaScience vol. 4 Preprint at https://doi.org/10.1186/s13742-015-0047-8 (2015).

71. Choi, S. W., Mak, T. S.-H. & O’Reilly, P. F. Tutorial: a guide to performing polygenic risk score analyses. Nat. Protoc. 15, 2759–2772 (2020).

72. Hinrichs, A. S. The UCSC Genome Browser Database: update 2006. Nucleic Acids Research vol. 34 D590–D598 Preprint at https://doi.org/10.1093/nar/gkj144 (2006).

