## Supplementary_information for "A comprehensive workflow for target adaptive sampling long-read sequencing applied to hereditary cancer patient genomes"

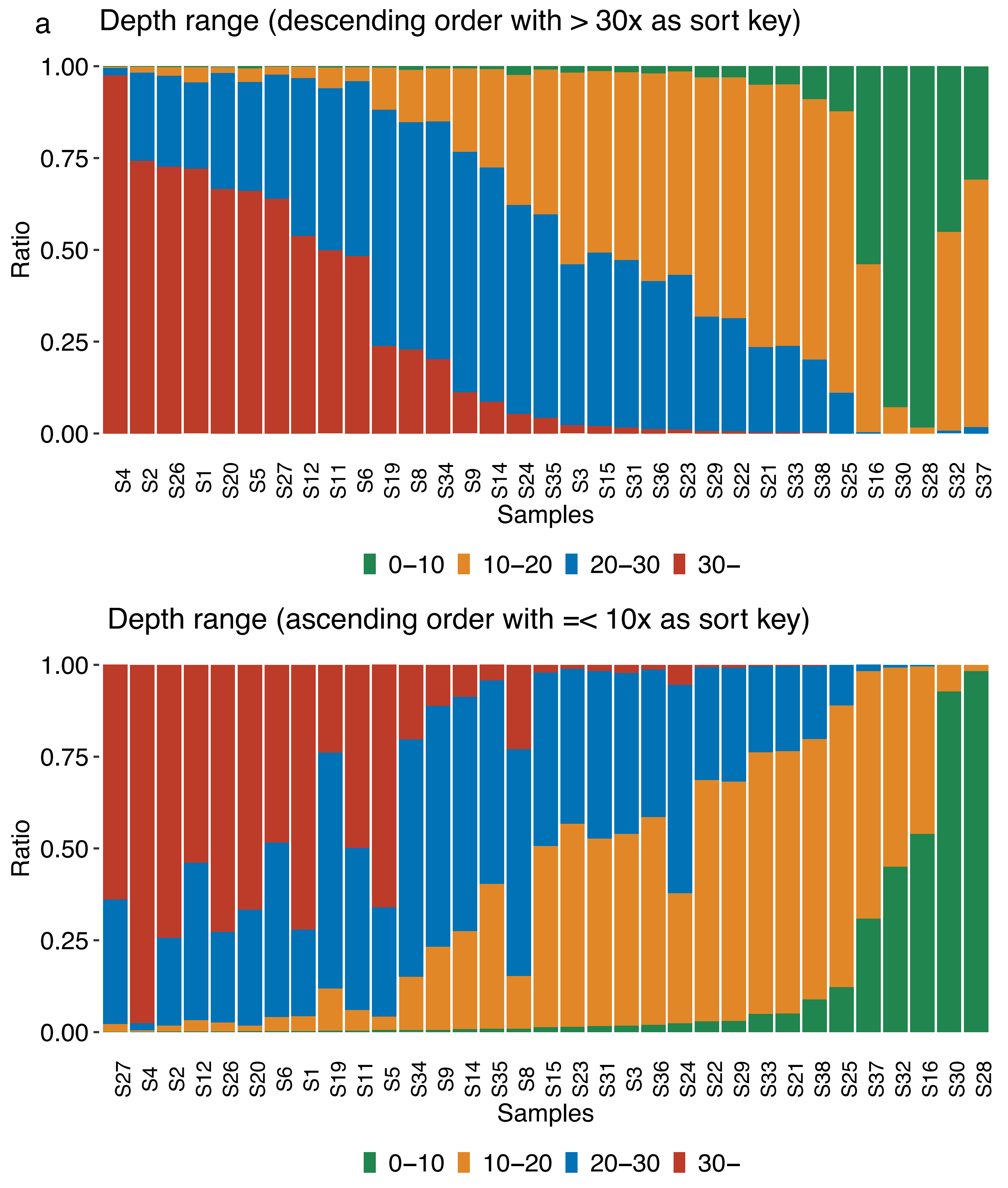


**Supplementary Figure 1**: **Summary of sequence coverage of TAS-LRS sequencing data.** The barplot shows the percentage of each sequence coverage range ([0, 10], [10, 20], [20, 30], [30, ∞] in the target area (a) in descending order with [30, ∞], and (b) in ascending order with [0, 10] as the sort keys.

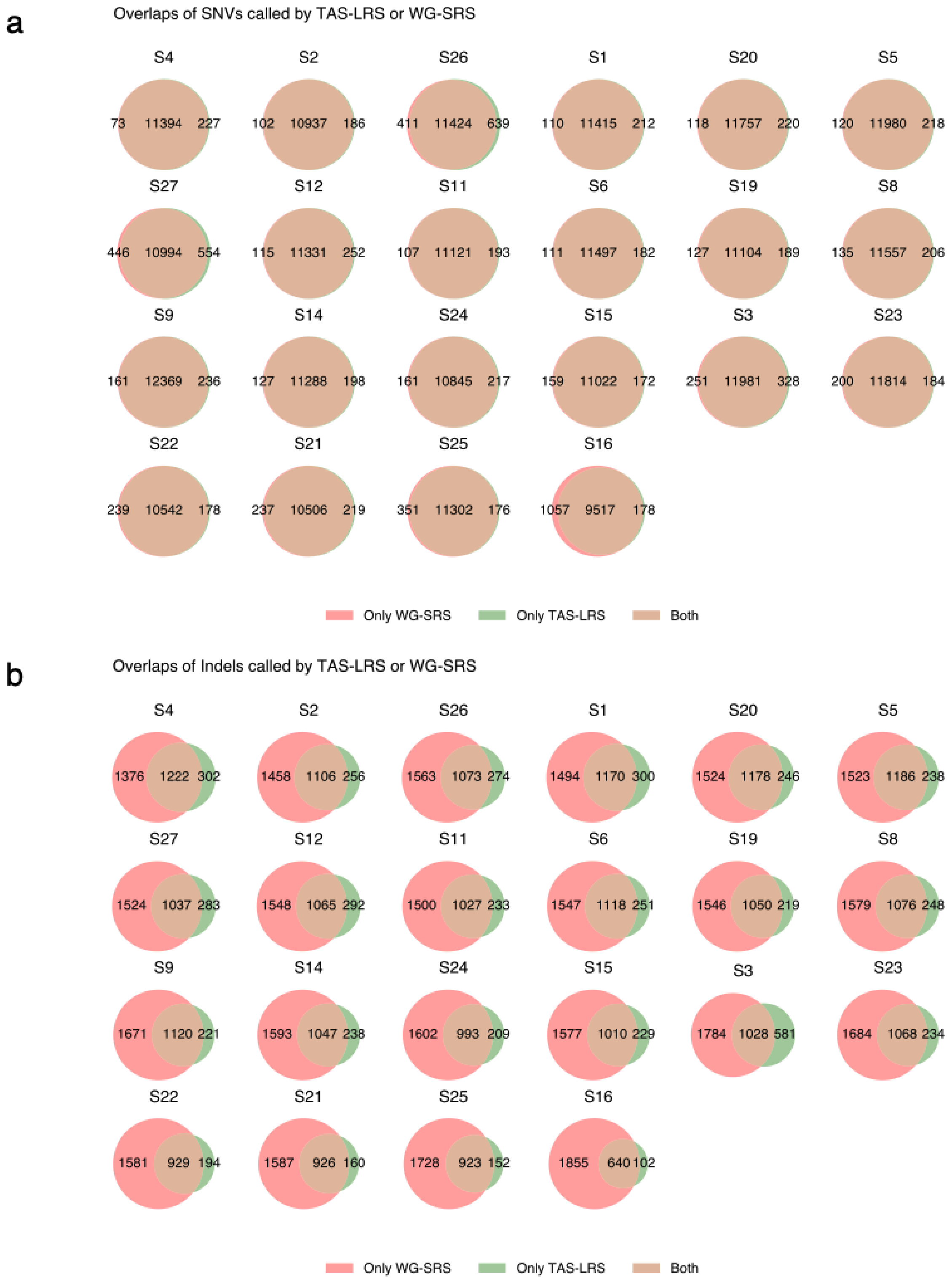


**Supplementary Figure 2: Venn diagrams of SNVs/Indels by TAS-LRS and WG-SRS for each sample**. Venn diagrams are sorted in descending order with mean sequence coverage in the target regions.

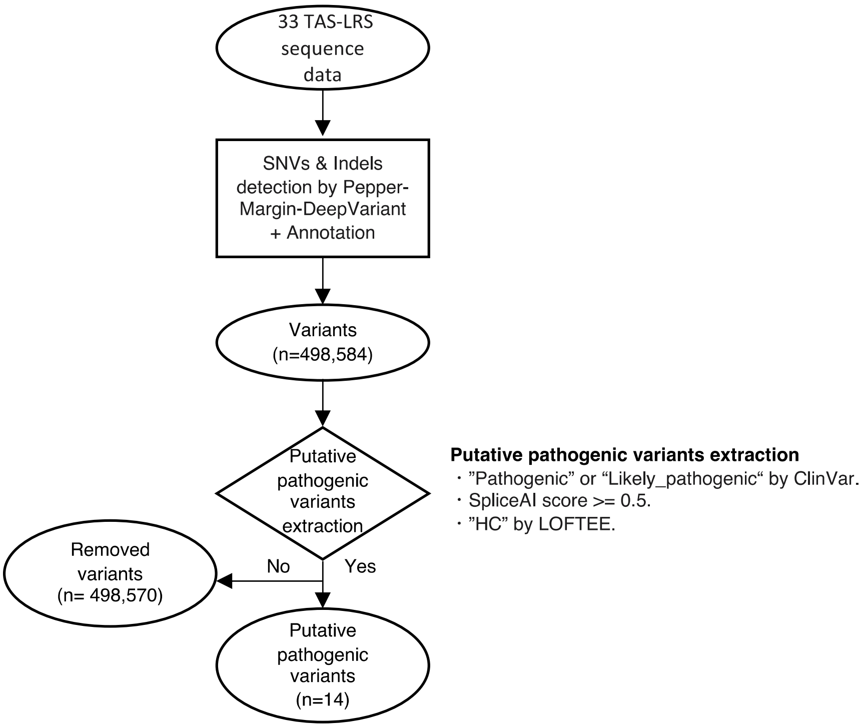


**Supplementary Figure 3: Flowchart for putative pathogenic SNVs/indels identification.**

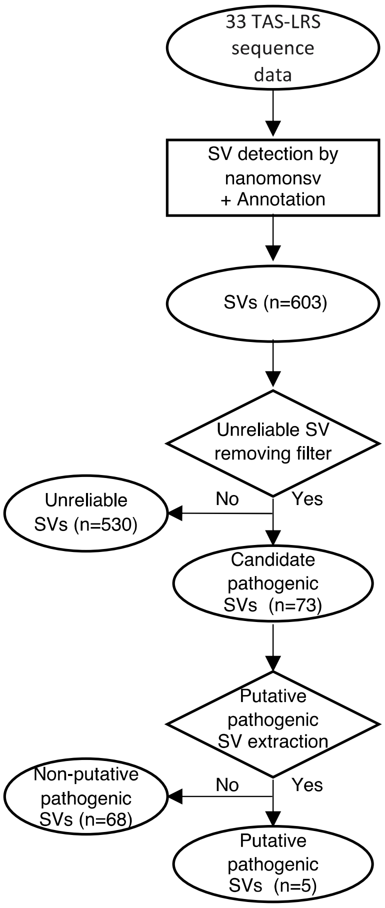


**Unreliable SV removing filter**

・One or both of the SV breakpoints are present in the unplaced contig, decoy sequence.
・Neither SV breakpoint is located within the target region.
・Deletion, insertion, and tandem duplication type SVs included in the simple repeat region.

**Putative pathogenic SV extraction**

・Deletions involving the coding regions of the 147 target genes.
・Duplications that alter the coding sequences of the 147 target genes.
・Inversions or translocations that disrupt the coding sequences of the 147 target genes.

Inversion breakpoint (n=2)：a patient with FAP (*APC*)
Translocation breakpoint(n=2)：a patient with RB (*RB1*)
sub-complete deletion with exon(n=1)：a patient with RB (RB1)

**Supplementary Figure 4: Flowchart for putative pathogenic SV detection for nanomonsv canonical SV module.**

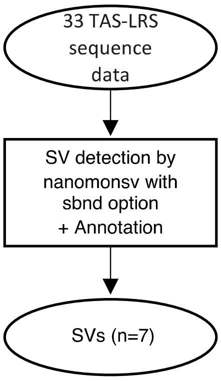


**Supplementary Figure 5: Flowchart for putative pathogenic SV detection for nanomonsv single-breakend SV module.**

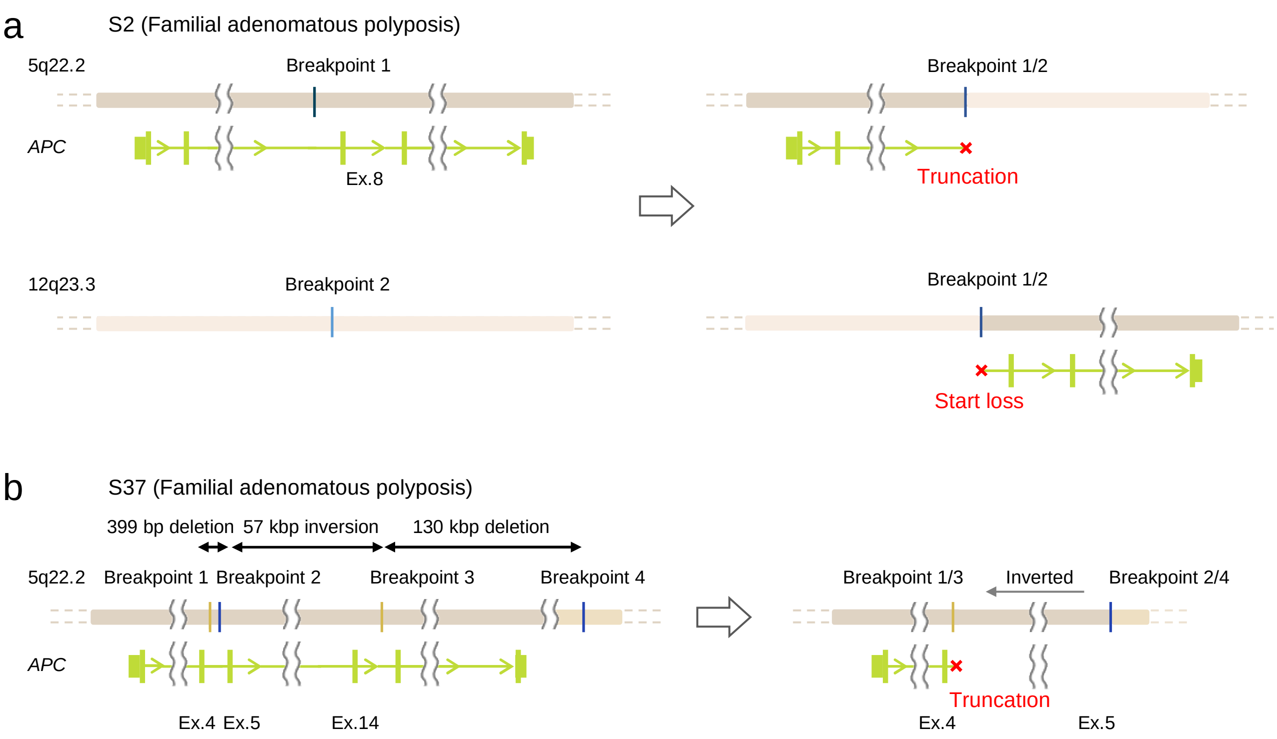


**Supplementary Figure 6: Schematic representation of structural variations of the *APC* gene in two patients with familial adenomatous polyposis.** (a) DNA swapping occurred at two breakpoints in the 7th intron of the *APC* gene and in the intergenic region at 12q23.3, forming a balanced translocation. (b) Reciprocal inversions occurred at breakpoints in the 4th and 14th intron of the *APC* gene, involving the deletions of 399 bp and 130 kbp.

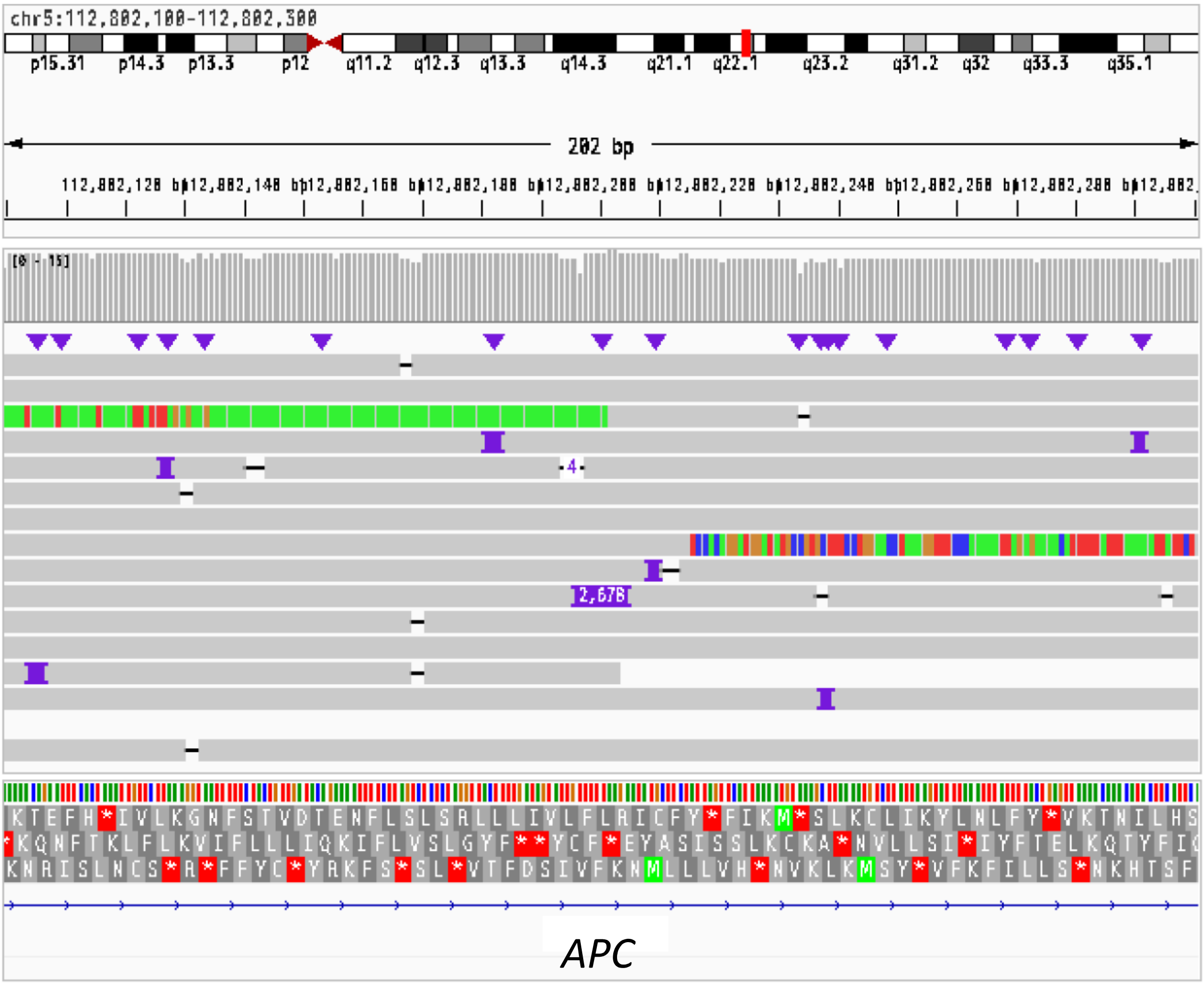


**Supplementary Figure 7: Details of SVA-derived insertion into the intronic region of the *APC* gene in a patient with familial adenomatous polyposis.** The IGV displayed long-read sequencing data and transcript sequencing data showing an SVA-derived insertion of 2,678 bp in the 8th intron of the *APC* gene.

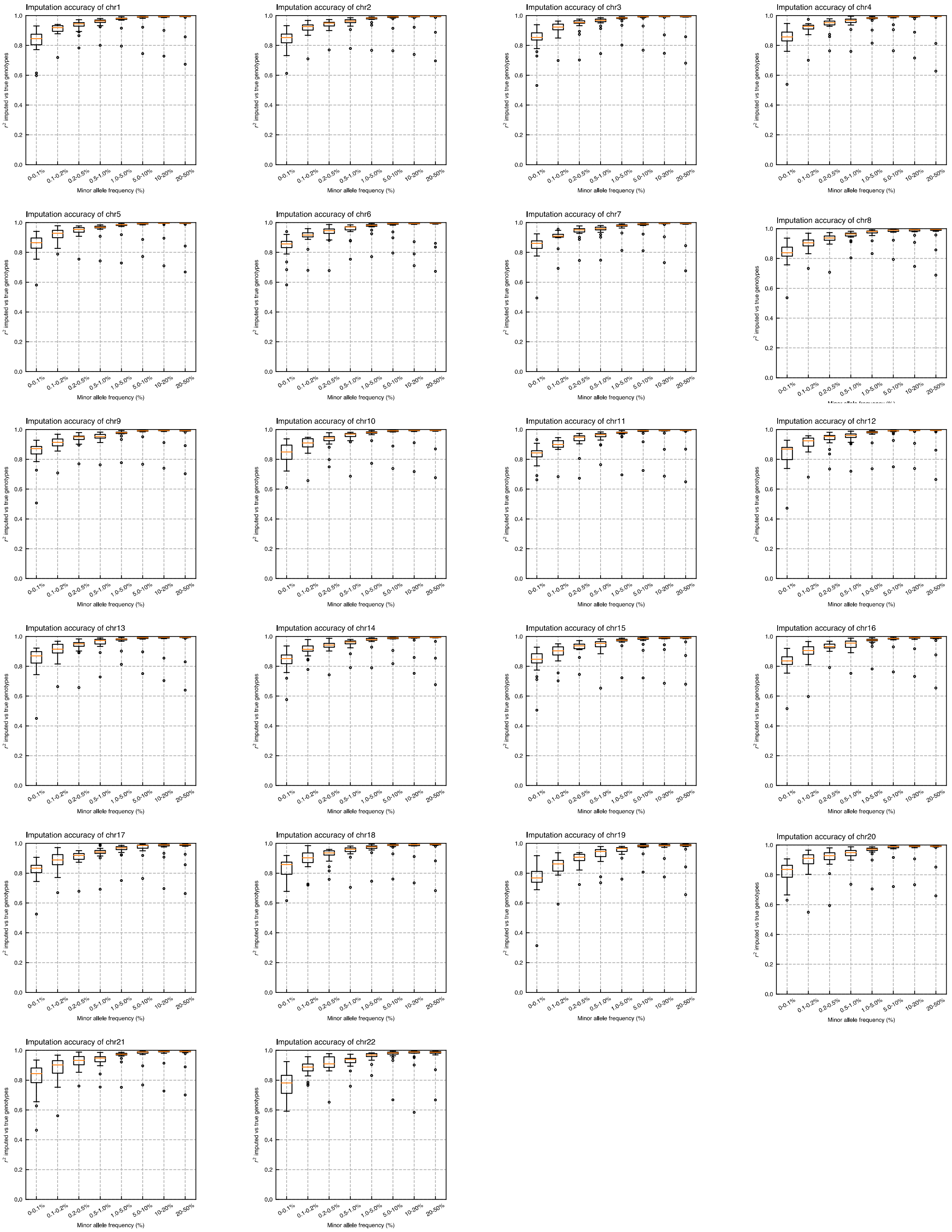


**Supplementary Figure 8: Imputation accuracy of TAS-LRS measured for each chromosome and each minor allele frequency range.** Genotyping by WG-SRS was used as the golden standard.

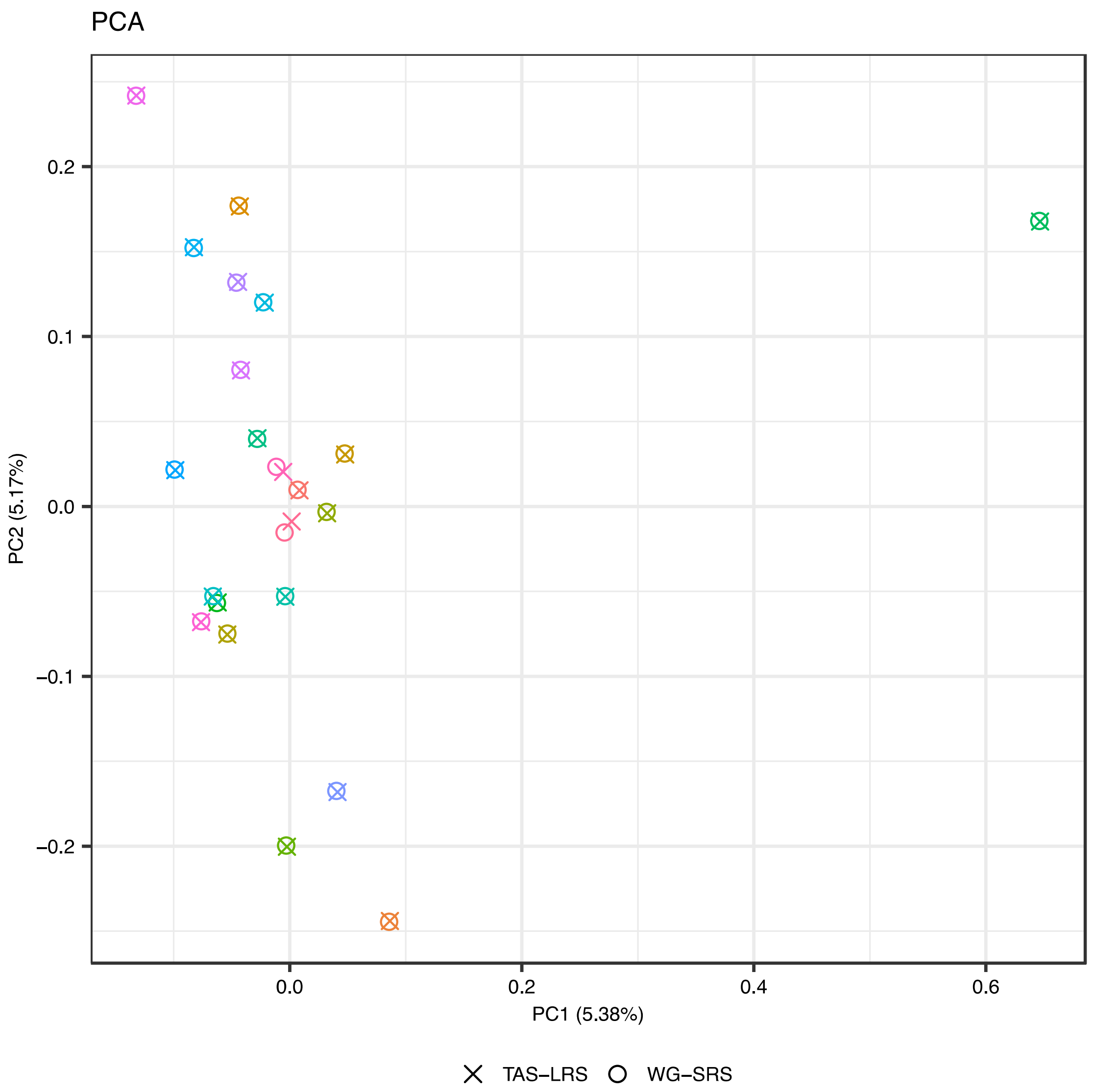


**Supplementary Figure 9**: **Principal component analysis (PCA) of genotype results from both TAS-LRS and WG-SRS for each individual (distinguished by color).** This figure includes one outlier sample probably originating from different ancestries. See also Figure 6b.

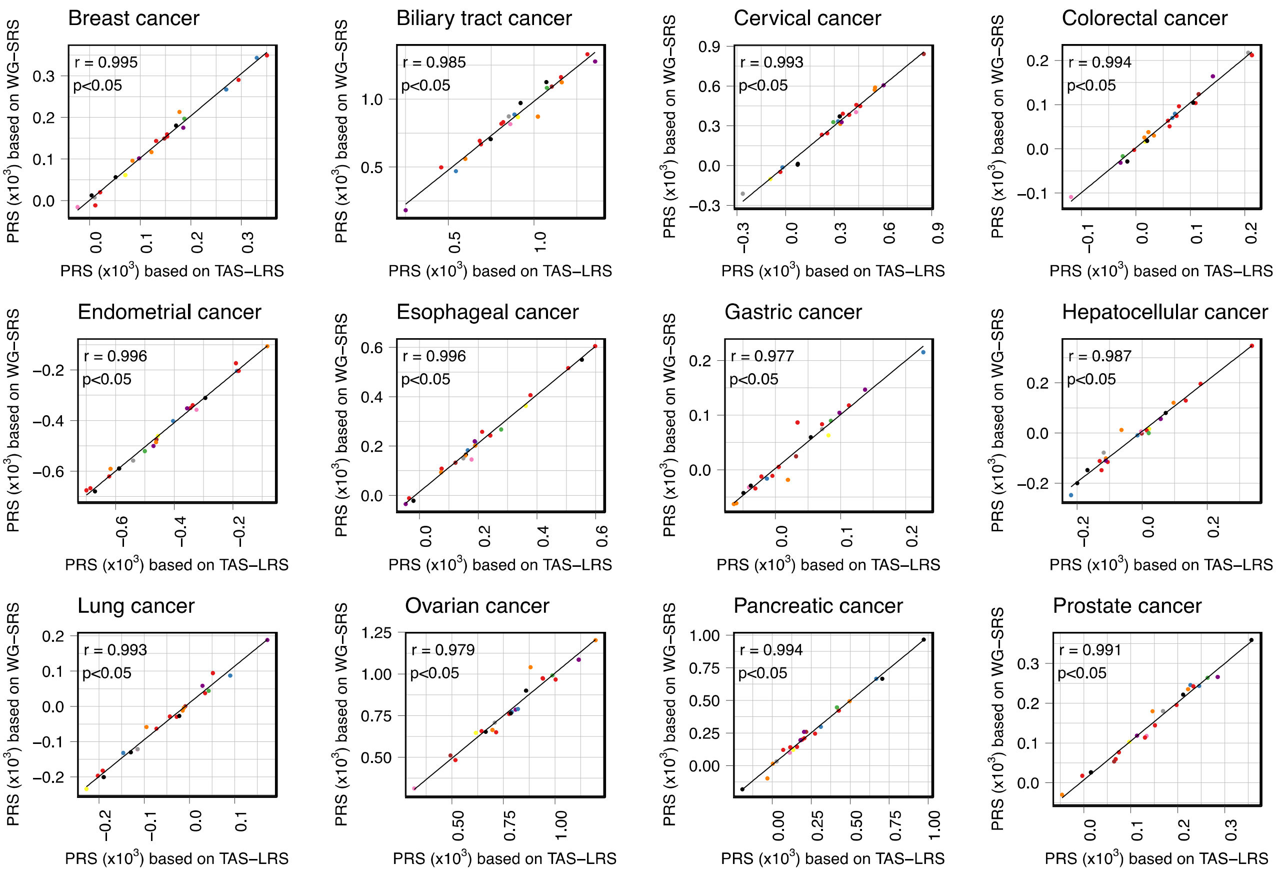


**Supplementary Figure 10**: **Comparison of Polygenic Risk Scores (PRSs) for 12 cancers calculated from the genotype by TAS-LRS (X-axis) and WG-SRS (Y-axis).** Each point indicates each sample and each color indicates each syndrome name (red: Familial adenomatous polyposis, blue: Familial pancreatic cancer, green: Hepatic angiomyolipoma, purple: Hereditary breast and ovarian cancer, orange: Li-Fraumeni syndrome, yellow: Lynch syndrome, brown: Multiple endocrine neoplasia type 1, pink: Multiple endocrine neoplasia type 2, gray: PTEN hamartoma tumor syndrome, black: Retinoblastoma).

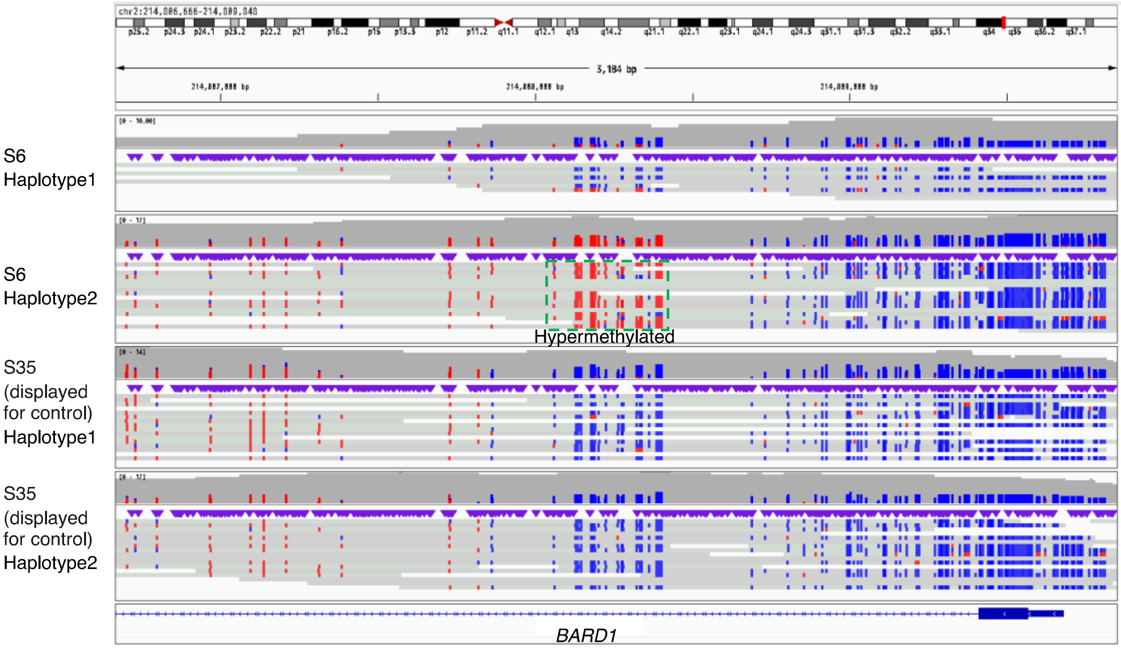


**Supplementary Figure 11: A case of a possible *BARD1* epimutation.** Alignment view of around the promoter region of the *BARD1* gene of the patient S3 and randomly selected control (S35). Each read was classified as haplotype 1 or 2 using Whatshap software. The CpG sites of each read are colored red if methylated and blue if not. It can be clearly seen that methylation is increased specifically for haplotype 2.
