## Supplementary_data_5 for "A comprehensive workflow for target adaptive sampling long-read sequencing applied to hereditary cancer patient genomes"

### NCBN Controls WGS Consortium

Hatsue Ishibashi-Ueda[1],Tsutomu Tomita[1], Michio Noguchi[1], Ayako Takahashi[1], Yu-ichi Goto[2], Sumiko Yoshida[3], Kotaro Hattori[3], Ryo Matsumura[3], Aritoshi Iida[4], Yutaka Maruoka[5], Hiroyuki Gatanaga[6], Masaya Sugiyama[7], Satoshi Suzuki[5], Kengo Miyo[8], Yoichi Matsubara[9], Akihiro Umezawa[10], Kenichiro Hata[11], Tadashi Kaname[12], Kouichi Ozaki[13], Haruhiko Tokuda[13], Hiroshi Watanabe[13], Shumpei Niida[13], Eisei Noiri[14], Koji Kitajima[14], Yosuke Omae[14,15], Reiko Miyahara[14], Hideyuki Shimanuki[14], Yosuke Kawai[15], and Katsushi Tokunaga[14,15]

[1] NCVC Biobank, National Cerebral and Cardiovascular Center, Suita, Osaka 564-8565, Japan
[2] Medical Genome Center, National Center of Neurology and Psychiatry, Kodaira, Tokyo 187-8551, Japan
[3] Department of Bioresources, Medical Genome Center, National Center of Neurology and Psychiatry, Kodaira, Tokyo 187-8551, Japan
[4] Department of Clinical Genome Analysis, Medical Genome Center, National Center of Neurology and Psychiatry, Kodaira, Tokyo 187-8551, Japan
[5] NCGM Biobank, National Center for Global Health and Medicine, Shinjuku-ku, Tokyo 162-8655, Japan
[6] AIDS Clinical Center, National Center for Global Health and Medicine, Shinjuku-ku, Tokyo 162-8655, Japan
[7] Genome Medical Sciences Project (Konodai), Research Institute, National Center for Global Health and Medicine, Ichikawa, Chiba 272-8516, Japan
[8] Center for Medical Informatics and Intelligence, National Center for Global Health and Medicine, Shinjuku-ku, Tokyo 162-8655, Japan
[9] National Center for Child Health and Development, Setagaya-ku, Tokyo 157-8535, Japan
[10] Center for Regenerative Medicine, National Center for Child Health and Development, Setagaya-ku, Tokyo 157-8535, Japan
[11] Department of Maternal-Fetal Biology, National Center for Child Health and Development, Setagaya-ku, Tokyo 157-8535, Japan
[12] Department of Genome Medicine, National Center for Child Health and Development, Setagaya-ku, Tokyo 157-8535, Japan
[13] Medical Genome Center, Research Institute, National Center for Geriatrics and Gerontology, Obu, Aichi 474-8511, Japan
[14] Central Biobank, National Center Biobank Network, Shinjuku-ku, Tokyo 162-8655, Japan
[15] Genome Medical Science Project (Toyama), Research Institute, National Center for Global Health and Medicine, Shinjuku-ku, Tokyo 162-8655, Japan
